# *BLOC1S1* variants cause lysosomal and autophagic defects resulting in a hypomyelinating leukodystrophy with epileptic encephalopathy

**DOI:** 10.1101/2025.07.17.25331211

**Authors:** Raffaella De Pace, Carlos Dominguez Gonzalez, Chad D. Williamson, Guy Helman, Leslie E. Sanderson, Brianna Disanza, Nicole Hsiao-Sánchez, Amy Pizzino, Kayla Muirhead, Joshua L Bonkowsky, Ryan J. Taft, Nouriya A Sannaa, Patricia Dias, Ana Sofia Quintas, Mehmet Burak Mutlu, Hasan Bas, Hasan Oztürk, Majid Mojarrad, Masoome Alerasool, Shahriar Sheikhani, Hayder Kadhim Jabbar, Awatif Hameed Issa, Henry Houlden, Emir Zonic, Tahsin Stefan Barakat, Kornelia Tripolski, Antonio Romito, Eden Teferedegn, Arastoo Vossough, Matthew T. Whitehead, Elizabeth Bhoj, Rebecca C. Ahrens-Nicklas, Cas Simons, Ernst Wolvetang, Tjakko J. van Ham, Aida M. Bertoli-Avella, Reza Maroofian, Juan S. Bonifacino, Adeline Vanderver

## Abstract

*BLOC1S1* encodes a subunit shared by the BLOC-1 and BORC hetero-octameric complexes that regulate various endolysosomal processes. Here, we report the identification of seven distinct variants in *BLOC1S1* in eleven individuals from seven independent families presenting with early psychomotor delay, hypotonia, spasticity, epileptic encephalopathy, optic atrophy, and leuko-axonopathy with hypomyelination. A subset of the affected individuals also have features of hypopigmentation and ocular albinism that are similar, although milder, than those of individuals with BLOC-1-related Hermansky-Pudlak syndrome. Functional analyses show that *BLOC1S1* knockout (KO) impairs the anterograde transport of lysosomes and autophagy in both non-neuronal cells and iPSC-derived neurons. Rescue experiments reveal that most *BLOC1S1* variants exhibit reduced expression, decreased assembly with other BORC/BLOC-1 subunits, and/or impaired restoration of lysosome transport and autophagy in *BLOC1S1*-KO cells. Additionally, we show that KO of *BLOC1S1* reduces pigmentation in a melanocytic cell line, and that five of the *BLOC1S1* variants partially or fully restore pigmentation. These findings provide genetic, clinical, and functional evidence that loss-of-function (LoF) of *BLOC1S1* leads to more pronounced deficits in BORC than BLOC-1 function. We conclude that the biallelic *BLOC1S1* variants characterized here primarily result in a neurological disorder with prominent leukodystrophy, similar to the recently reported condition caused by variants in the BORCS8 subunit of BORC. Together, these findings establish BORCopathies as a distinct disease entity.

## Introduction

The Biogenesis of Lysosome-related Organelles Complex-1 (BLOC-1)^1,2,3^ and BLOC-One-Related Complex (BORC)^4^ are structurally related eight-subunit complexes that regulate various endolysosomal processes.^5^ BLOC-1 is involved in the biogenesis of lysosome-related organelles (LROs)^6,7^ such as melanosomes in melanocytes and platelet dense bodies in megakaryocytes,^8,9,10,11^ and in the formation of endosomal recycling tubules in non-specialized cells.^12,13,14,15^ In contrast, BORC promotes ARL8-dependent coupling of lysosomes to kinesin motors for anterograde transport along microtubules^4,16,17,18,19^ and to the HOPS complex for fusion of lysosomes to late endosomes and autophagosomes.^20,21,22^ BLOC-1 and BORC each consist of five complex-specific subunits, with three subunits shared between the two complexes (Table S1).

Biallelic loss-of-function (LoF) variants in four genes encoding BLOC-1-specific subunits are associated with Hermansky-Pudlak syndrome (HPS) (MIM:614077), a disorder characterized by oculocutaneous albinism, platelet dysfunction, and visual defects,^23^ while biallelic LoF variants in one of the five BORC-specific subunits is associated with an early-onset neurodegenerative disorder characterized by global developmental delay, hypotonia, spasticity, muscle wasting, dysmorphic facies, optic atrophy, and leuko-axonopathy with hypomyelination^19^ (Table S1).

Additionally, biallelic variants in two other BORC-subunit genes, *BLOC1S1* (OMIM:601444)^24^ and *BORCS5* (OMIM:616598),^25^ were identified in large genomic cohort studies of patients with neurological disorders, although the clinical features were not characterized in detail and the functional properties of the variant proteins were not assessed. These findings suggest that disease phenotypes segregate distinctly with the affected complex, with LoF variants in BLOC-1-specific subunits resulting in HPS while LoF variants in BORC specific-subunits result in primary neurological disorders.

Here, in addition to the originally reported four individuals from three independent families with biallelic variants in *BLOC1S1,*^24^ we have now identified seven additional affected individuals from five other independent families. All individuals harboring *BLOC1S1* variants with available neuroimaging exhibit hypomyelinating leukodystrophy characteristic of BORC deficiency.^19^

## Subjects and methods

### Subject ascertainment and clinical and molecular studies

Seven unrelated families of six different ancestry backgrounds were recruited and studied (Figure 1A, Tables 1 and S2). The study was approved by the institutional ethics committees of the Children’s Hospital of Philadelphia (IRB# 14-011236), University College London (IRB# 310045/1571740/37/598), and CENTOGENE, as previously described.^24^ Written informed consent to site-specific protocols was obtained from all seven families in accordance with the Declaration of Helsinki.

**Figure 1.**
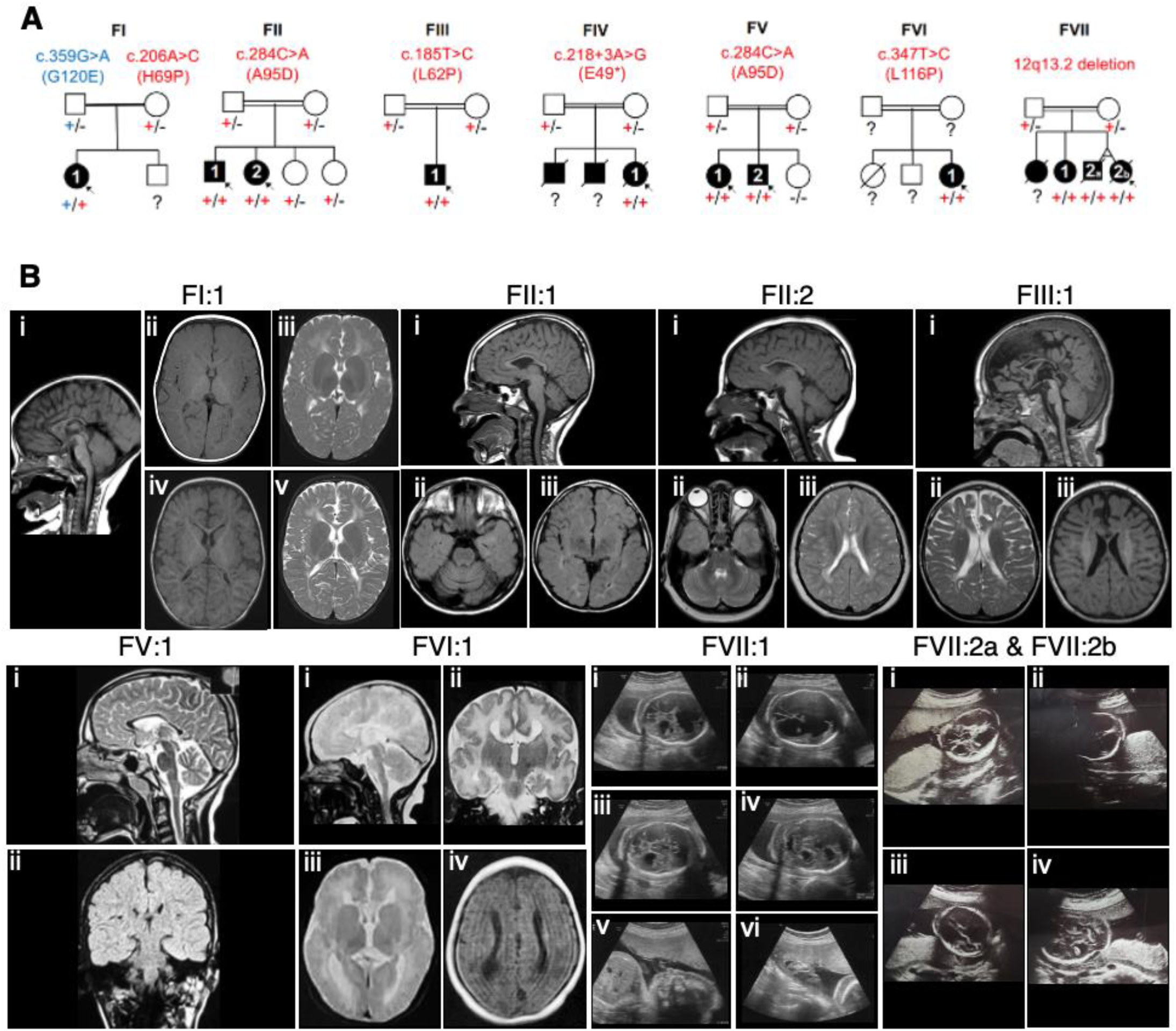
Clinical pedigrees and neuroimaging features of *BLOC1S1*-associated hypomyelinating leukodystrophy with epileptic encephalopathy **(A)** Pedigrees showing segregation of variants (+) in the *BLOC1S1* gene. In individual FI:1 there are two heterozygous variants in trans. In the remaining individuals (FII:1, FII:2, FIII:1, FIV:1, FV:1, FV:2, FVI:1, FVII:1, FVII:2a, FVII:2b) the variants are homozygous. **(B)** Brain MRI images corresponding to individuals FI:1, FII:1, FII:2, FIII:1, FV:1, FVI:1, and prenatal ultrasound images corresponding to Individuals FVII:1, FVII:2a, FVII:2b. MRI was not performed or imaging was not available for individuals FIV:1, FV:2, FVII:1. Brain MR images show variable hypomyelination with or without superimposed sequela of deep cerebral white matter dysmyelination or demyelination, corpus callosum abnormalities (hypoplasia, hypogenesis/dysgenesis, or agenesis), and brain volume loss. Fetal ultrsound images demonstrate ventriculomegaly and midline abnormalites. See Supplemental information for detailed image descriptions.

Detailed clinical features, brain MRI imaging (Figure 1B), family history and clinical notes were obtained from affected individuals and reviewed by a group of clinical geneticists and pediatric neurologists. Brain MRIs were reviewed by two experienced pediatric neuroradiologists. Genome, exome, and Sanger sequencing were performed independently at different research and clinical laboratories, as described previously.^24,26^

Another individual was initially suspected to have a *BLOC1S1*-related condition after biallelic variants in *BLOC1S1* were identified [c.343G>T, p.Ala115Ser (A115S)]. However, there was lack of overlap in clinical phenotype. Functional studies performed independently of clinical and genetic phenotyping indicated normal BLOC1S1 function. Subsequently, this variant was considered not clinically relevant and the sample used as control (see Control in Table 1).

**Table 1.**
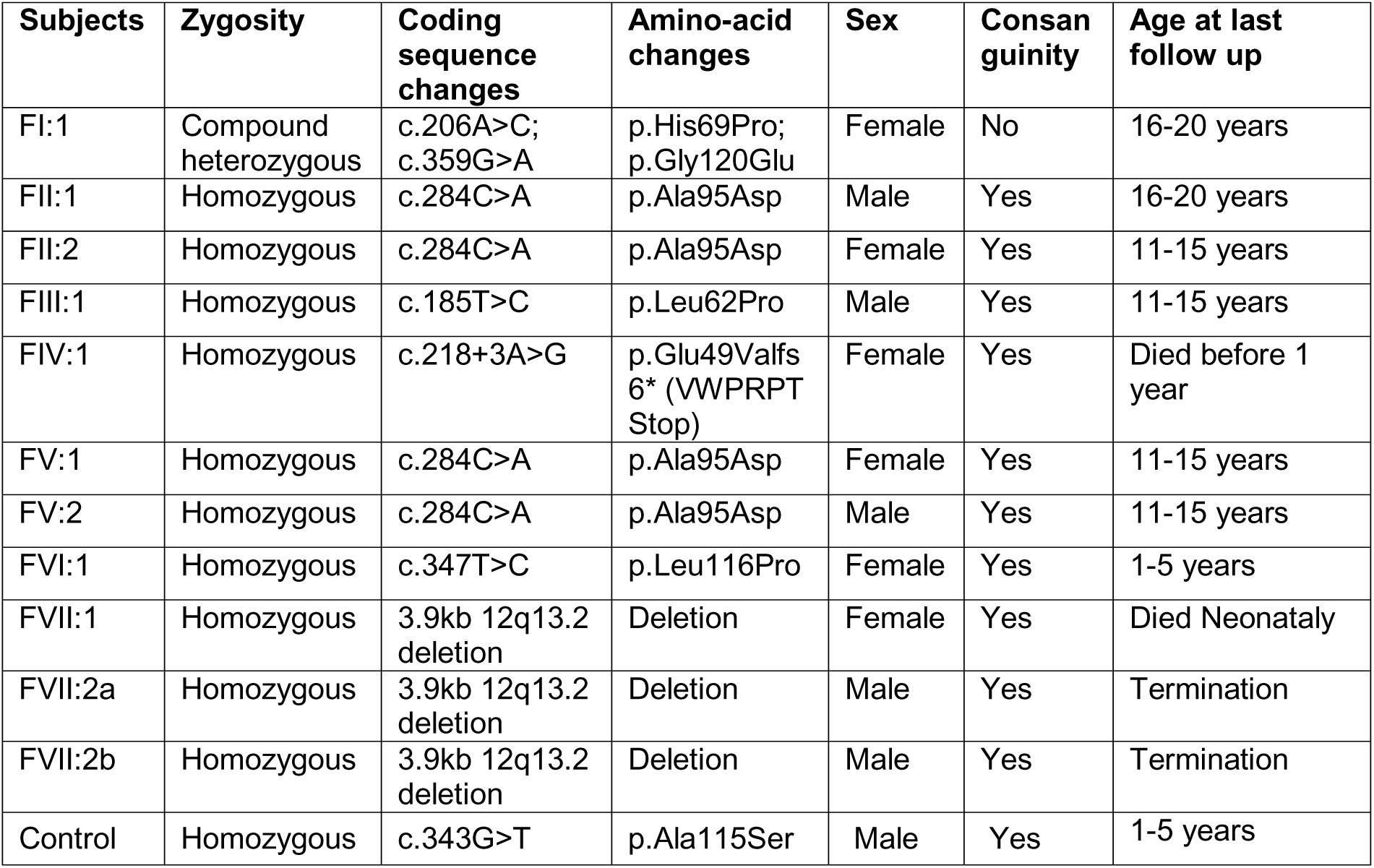
Summary of genotypes and demographics.

### In silico characterization of *BLOC1S1* variants

To compare amino-acid sequence conservation of human BLOC1S1 (Figure 2A) to orthologs from other species, protein sequences from *Homo sapiens* (NP_001478.2), *Pan troglodytes* (XP_509120.1), *Bos taurus* (XP_059742705.1), *Sus scrofa* (XP_003126301.3), *Gallus gallus* (XP_015128068.1), *Mus musculus* (NP_056555.2), *Xenopus tropicalis* (NP_001072249.1), *Danio rerio* (NP_001037815.1), *Drosophila melanogaster* (NP_725401.2), and *Caenorhabditis elegans* (NP_499262.2) were aligned using ClustalO multiple sequence alignment, version 1.2.4 (https://toolkit.tuebingen.mpg.de/tools/clustalo). For each residue of human BLOC1S1, the corresponding residue (if any) in each orthologous protein was found from the ClustalO alignment and given a value of 1 if it exactly matched the human sequence, 0.5 if it was a conservative substitution, or 0 if it was a non-conservative substitution or was absent. Total scores for each residue were determined and plotted using a color scale to differentiate regions of variable sequence conservation (Figure 2B).

**Figure 2.**
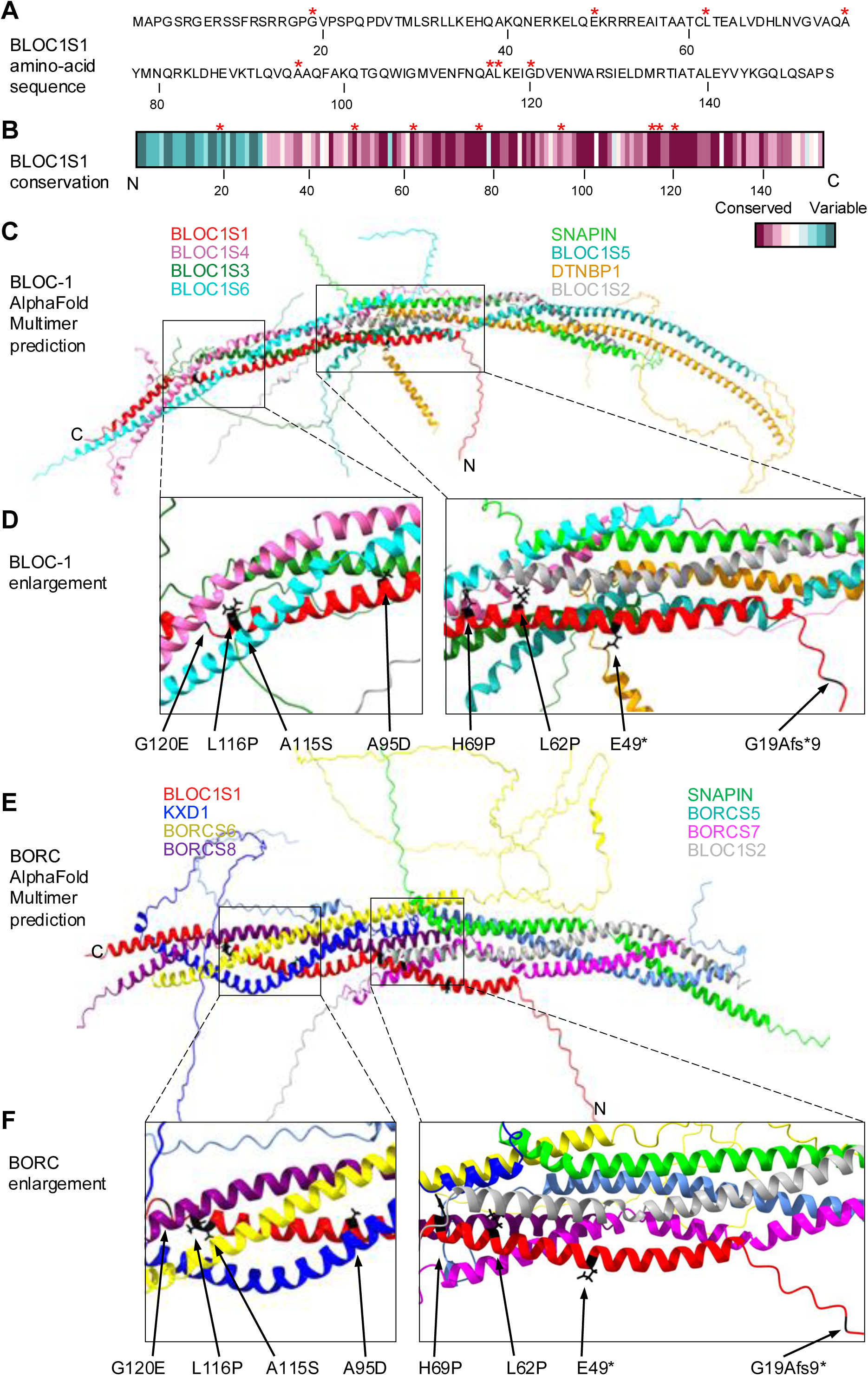
Predicted structural features of BLOC-1 and BORC, and variant locations **(A)** Amino-acid sequence of WT human BLOC1S1 (NP_001478.2) indicating the positions of the variants (red asterisks). **(B)** Amino-acid sequences of BLOC1S1 from 9 different species aligned to human BLOC1S1. Sequence conservation was calculated as a percent residue match between species with the human sequence, as described in the Materials and methods section, and represented graphically with a blue (low conservation between species) to maroon (high conservation between species) scale. **(C)** Structure of BLOC-1 predicted by AlphaFold Multimer. BLOC-1 subunits are shown in different colors. N- and C-termini of BLOC1S1 are indicated. **(D)** Close-up views of the positions of BLOC1S1 variants in BLOC-1 (highlighted in black). **(E)** Structure of BORC predicted by AlphaFold Multimer. BORC subunits are shown in different colors. N- and C-termini of BLOC1S1 are indicated. **(F)** Close-up views of the positions of BLOC1S1 variants in BORC (highlighted in black).

BLOC-1 (composed of BLOC1S1 [NP_001478.2], BLOC1S2 [NP_776170.2], SNAPIN [NP_036569.1], BLOC1S3 [NP_997715.1], BLOC1S4 [NP_060836.1], BLOC1S5 [NP_958437.1], BLOC1S6 [NP_036520.1], and DTNBP1 [NP_115498.2]), and BORC (composed of BLOC1S1 [NP_001478.2], BLOC1S2 [NP_776170.2], SNAPIN [NP_036569.], KXD1 [NP_001165419.1], BORCS5 [NP_477517.1], BORCS6 [NP_060092.2], BORCS7 [NP_653192.2], and BORCS8 [NP_001139256.1]) were modeled (Figure 2C-F) using the AlphaFold2 version 2.3.2 build on the NIH HPC Biowulf cluster, with a database max template date set to 2024-01-20. Highest ranked models were analyzed in ChimeraX (Figure 2C and E). Protein-protein interactions were determined from predicted contacts within a distance of 4 Angstroms (Figure S1). SpliceAI was used to predict the impact of the *BLOC1S1* variants on transcript splicing and translation.

### Recombinant DNAs and mutagenesis

For transfection of HEK293T and HeLa cells, the human *BLOC1S1* cDNA with sequences encoding 13 copies of the myc-epitope tag at the C-terminus was cloned using Gibson assembly (New England Biolabs) into a pCI-neo plasmid (Promega). The shorter protein (AAB37682.1) of 125 amino acids was used for the cloning. The 29 amino acids missing in this protein are predicted to be disordered and have the lowest conservation score amongst homologs because they are absent from the protein in catalogued sequences of most species (Figure 2B, C and E). For mutagenesis, we used the QuikChange II XL Site-Directed Mutagenesis Kit (Agilent) according to the manufacturer’s instructions. To introduce the *BLOC1S1* variants in the WT construct, we designed one forward and one reverse oligonucleotide bearing the selected variant (lowercase): E49*, forward 5′-CAAGCAGAATGAACGCAAGGAGCTGCAGgtgtggcctaggcctacaGTCGACCCCGGGATAATTAACGGTGAAC and reverse 5′-GTTCACCGTTAATTATCCCGGGGTCGACtgtaggcctaggccacacCTGCAGCTCCTTGCGTTCAT TCTGCTTG. L62P, forward 5′-CTGCAGCGACCTGCCcGACAGAAGCTTTGGT and reverse 5′-ACCAAAGCTTCTGTCgGGCAGGTCGCTGCAG. H69P, forward 5′-AAGCTTTGGTGGATCcCCTCAATGTGGGTGT and reverse 5′-ACACCCACATTGAGGgGATCCACCAAAGCTT. A95D, forward 5′-CCCTACAGGTCCAGGaTGCCCAATTTGCCAA and reverse 5′-TTGGCAAATTGGGCAtCCTGGACCTGTAGGG. A115S, forward 5′-AGAACTTCAACCAGtCACTCAAGGAAATTG and reverse 5′-CAATTTCCTTGAGTGaCTGGTTGAAGTTCT. G120E, forward 5′-CACTCAAGGAAATTGaGGATGTGGAGAACTG and reverse 5′-CAGTTCTCCACATCCtCAATTTCCTTGAGTG. Constructs were Sanger-sequenced to verify the changed base pairs in the coding sequence.

For transduction of MNT-1 (human malignant melanoma line) cells, the human *BLOC1S1* cDNA was cloned into a pLV[Exp] lentiviral plasmid (VectorBuilder) with sequences encoding one copy of the HA-epitope tag at the C-terminus and an EF1α promoter. The plasmid also encodes enhanced GFP (EGFP) under the control of the CMV promoter to allow the selection of infected cells. The canonical isoform (NP_001478.2) of 153 amino acids was used for the cloning. To introduce the *BLOC1S1* variants in the WT construct, we used the QuikChange II XL Site-Directed Mutagenesis Kit (Agilent) according to the manufacturer’s instructions. For the E49* and G120E variants, we used primers to create blunt-end PCR products using Q5 High-Fidelity 2X Master Mix (Biolabs) that were then joined by KLD Enzyme Mix 5 min reaction (Biolabs). For each variant, we designed one forward and one reverse oligonucleotide: E49*, forward 5′-ctaggcctacaTACCCATACGATGTTCCAG and reverse 5′-gccacacCTGCAGCTCCTTGCGTTCATTC. L62P, forward 5′-CTGCAGCGACCTGCCcGACAGAAGCTTTGGT and reverse 5′-ACCAAAGCTTCTGTCgGGCAGGTCGCTGCAG. H69P, forward 5′-AAGCTTTGGTGGATCcCCTCAATGTGGGTGT and reverse 5′-ACACCCACATTGAGGgGATCCACCAAAGCTT. G120E, forward 5′-TTGaGGATGTGGAGAACTG and reverse 5′-TTTCCTTGAGTGCCTGGTT. A95D, forward 5′-CCCTACAGGTCCAGGaTGCCCAATTTGCCAA and reverse 5′-TTGGCAAATTGGGCAtCCTGGACCTGTAGGG. A115S, forward 5′-AGAACTTCAACCAGtCACTCAAGGAAATTG and reverse 5′-CAATTTCCTTGAGTGaCTGGTTGAAGTTCT. Constructs were Sanger-sequenced to verify the changed base pairs in the coding sequence.

### Non-neuronal cell culture and transfection

HeLa and HEK293T cells (ATCC) were cultured in Dulbecco’s Modified Eagle’s Medium (DMEM) (Quality Biological), supplemented with 2 mM L-glutamine (GIBCO), 10 % fetal bovine serum (FBS) (Corning), 100 U/mL penicillin-streptomycin (GIBCO) (complete DMEM) in a 37 °C incubator at 5% CO_2_ and 95% air. HeLa cells grown on 6-well plates were transiently transfected with 2 μg of each plasmid DNA using 8 μL Lipofectamine 3000 (Invitrogen) according to the manufacturer’s protocols. Approximately 24 h after transfection, cells were split onto 12-mm coverslips coated with collagen (BioTechne). Cells were then cultured for an additional 24 h before fixation and immunofluorescent labeling. A 48 hour transfection period was necessary for rescue of the normal *BLOC1S1* phenotype. For the co-immunoprecipitation experiments, HEK293T cells grown on 10-cm plates were transiently transfected for 48 h with 10 μg plasmid DNA and 25 μl Lipofectamine 2000 (Invitrogen) according to the manufacturer’s instructions. One plate was used for the transfection of each construct in each experiment. To culture MNT-1 cells (ATCC), we used DMEM supplemented with 20% FBS, 10% AIM V (ThermoFisher Scientific), 1x non-essential amino acids (NEAA) (Thermo Fisher Scientific), 100 U/mL penicillin-streptomycin (GIBCO) in a 37 °C incubator at 5% CO_2_ and 95% air. *BLOC1S1*-KO MNT-1 cells grown on 6-well plates were infected with lentivirus for 48 h. Cells were then lifted with 0.25% trypsin-EDTA (GIBCO), and cells positive for GFP were sorted into a 6-well plate.

### CRISPR-Cas9 knockout (KO) of *BLOC1S1* and *BLOC1S6*

To KO the *BLOC1S1* gene in HeLa cells, MNT-1 cells and iPSCs, and *BLOC1S6* in HeLa cells, we used the CRISPR-Cas9 system with a single guide (sg) RNA per gene. For *BLOC1S1* KO, the sgRNA sequence was designed using Benchling (https://benchling.com) to target the first exon. We designed one forward and one reverse oligonucleotide bearing the selected sgRNA (uppercase) and the sequence necessary for cloning into the px458 plasmid (lowercase): forward 5′-caccgGGCCAAGCAGAATGAACGCA and reverse 5’-aaacTGCGTTCATTCTGCTTGGCCc. For *BLOC1S6* KO, we used Benchling to design oligonucleotides bearing an sgRNA sequence targeting exon 1: forward 5’-caccgACGGCCACCCTACTGCCTGG and reverse 5’-aaacCCAGGCAGTAGGGTGGCCGTc. Each of the guides was separately cloned into the px458 plasmid (gift from Feng Zhang, Addgene plasmid #48138). Briefly, the px458 plasmid was cut with BbsI and the annealed oligonucleotides were inserted using T4 DNA ligase. The insertion was verified by sequencing. Plasmids bearing the sgRNAs were co-transfected into HeLa and iPSCs using Lipofectamine 2000 (Thermo Fisher Scientific). Forty-eight hours later, single cells were sorted into 96-well plates based on GFP signal. The single cells were allowed to grow in complete E8-Flex medium supplemented with RevitaCell (Thermo Fisher Scientific) for iPSCs, and complete DMEM for HeLa cells. After 1-2 weeks, colonies were visible, and cells were transferred to a 6-well dish and grown to confluency. Cells were then harvested, and genomic DNA was isolated. PCR was conducted using forward 5’-AGCGGTCACGTGACAT and reverse 5’-AATAGGCAGGGCGCT primers for *BLOC1S1*, and forward 5’-CGTTTGCTTCTTCCCTGTGTT and reverse 5’-CGGAGTTCTACTCACTCAGG primers for *BLOC1S6*. The PCR products were cloned into pCR™-Blunt II-TOPO™ Vector (Thermo Fisher Scientific) and sequenced to verify the change from the WT sequence. Further verification of the BLOC1S1 absence was conducted by SDS-PAGE and immunoblotting.

### SDS-PAGE and immunoblotting

Cells were washed twice with ice-cold phosphate-buffered saline (PBS; Corning), scraped from the plates in 1x Laemmli sample buffer (1xLSB) (Bio-Rad) supplemented with 2.5% v/v 2-mercaptoethanol (Sigma-Aldrich), heated at 95 °C for 5 min, and resolved by SDS-PAGE. Gels were blotted onto nitrocellulose membranes and blocked with 5% w/v non-fat milk in Tris-buffered saline, 0.1% v/v Tween 20 (TBS-T) for 20 min. Membranes were sequentially incubated with primary antibody and secondary HRP-conjugated antibody diluted in TBS-T. SuperSignal West Dura Reagents (Thermo Fisher) were used for detection of the antibody signal with a Bio-Rad Chemidoc MP imaging system. Detection of β-tubulin or β-actin was used as a control for sample loading.

### Human *NGN2*-iPSC culture and neuronal differentiation

Human iPSCs expressing doxycycline-inducible *NGN2* (neurogenin)^27,28^ were cultured on Matrigel-coated dishes in Essential 8 Flex Medium (E8Flex) (Thermo Fisher Scientific). Cells were passaged with Accutase (Stem Cell Technologies) when 70% confluent, and seeded into E8Flex supplemented with 10 μM Y-27632 dihydrochloride ROCK inhibitor (Stem Cell Technologies). After 24 h, the medium was replaced with E8Flex without ROCK inhibitor and maintained until the next passage. For iPSCs differentiation into i3Neurons, iPSCs were seeded on day 0 in induction medium containing DMEM/F12 (Thermo Fisher Scientific) with 1x NEAA (Thermo Fisher Scientific), 1x GlutaMAX (Thermo Fisher Scientific), 1x N2A supplement (Thermo Fisher Scientific), 2 μg/mL doxycycline (Millipore Sigma), and 10 μM Y-27632 dihydrochloride ROCK inhibitor (Selleckchem). The induction medium was changed every day for three days. Cells were then detached with Accutase, counted and plated in neuronal culture medium consisting of BrainPhys medium (Stem Cell Technology) supplemented with 10 ng/mL neurotrophin-3 (Thermo Fisher Scientific), 10 ng/mL BDNF (Thermo Fisher Scientific), 1x B-27™ Supplement serum free (Thermo Fisher Scientific), 2 μg/mL doxycycline (Millipore Sigma), and 1 μg/mL mouse laminin (Thermo Fisher Scientific) (complete BrainPhys). For immunofluorescence microscopy of i3Neurons, the coverslips were pre-treated (etched) with 37% HCl, washed, autoclaved and coated with poly-L-lysine hydrobromide (overnight) (Millipore Sigma) and laminin for 1 h (Millipore Sigma). For SDS-PAGE of i3Neurons, the 12-well plate was coated with polylysine-laminin as for the coverslips. Every three days, half of the medium was removed, and an equal volume of fresh complete BrainPhys medium was added to maintain neuronal health.

### Immunofluorescence microscopy

HeLa cells were seeded on collagen-coated coverslips in 24-well plates at 40,000 cells per well in complete DMEM. i3Neurons were seeded on polylysine-laminin-coated 18-mm coverslips in 12-well plates at 50,000 cells per well and cultured in complete BrainPhys for 20 days. Cells were then fixed in 4% v/v paraformaldehyde (Electron Microscopy Sciences) in PBS for 20 min, permeabilized and blocked with 0.1% w/v saponin, 1% w/v BSA (Gold Bio) in PBS for 20 min, and sequentially incubated with primary and secondary antibodies diluted in 0.1% w/v saponin, 1% w/v BSA in PBS for 45 min at 37°C. Coverslips were washed three times in PBS and mounted on glass slides using Fluoromount-G (Electron Microscopy Sciences) with DAPI. Z-stack cell images were acquired on a Zeiss LSM 900 inverted confocal microscope (Carl Zeiss) using a Plan-Apochromat 63X oil objective (NA=1.4). Maximum intensity projections and final composite images were created using ImageJ/Fiji (https://fiji.sc/).

### Immunoprecipitation

Transfected HEK293T cells were lifted in ice-cold PBS and centrifuged at 500 x *g* for 5 min. The pellet was washed twice with ice-cold PBS and lysed in 10 mM Tris-HCl pH 7.5, 150 mM NaCl, 0.5 mM EDTA, 0.5% v/v Nonidet P40 supplemented with a protease inhibitor cocktail (Roche).

The cell lysate was centrifuged at 17,000 x *g* for 10 min. Ten percent of the supernatant was saved as input and the rest was incubated with anti-myc magnetic agarose beads (Thermo Fisher) at 4 °C for 1 h. After three washes with 10 mM Tris-HCl pH 7.5, 150 mM NaCl, 0.5 mM EDTA, the precipitates were eluted with 1x LSB at 95 °C for 5 min. The immunoprecipitated samples and inputs were analyzed by SDS-PAGE and immunoblotting.

### Antibodies and chemicals

The following primary antibodies were used for immunoblotting (IB) and/or immunofluorescence microscopy (IF): rabbit anti-BLOC1S1 (from Michael N. Sack’s laboratory, IB 1:1,000), rabbit anti-BLOC1S5 (24015-1-AP, Proteintech, IB 1:1,000), rabbit anti-BORCS5 (17169-1-AP, Proteintech, IB 1:1,000), rabbit anti-BORCS7 (PAB23142, Abnova, IB 1:1,000), rabbit anti-myc-tag (2272, Cell Signaling, IB 1:1,000), mouse anti-LAMP1 (H4A3, DSHB, IF 1:500), rabbit anti-LC3 (3868, Cell Signaling, IF 1:200, IB 1:1,000), mouse anti-α-tubulin-HRP (sc-32293, Santa Cruz, IF 1:1,000), mouse anti-β-actin-HRP (A3854, Millipore, IB 1:2,000), chicken anti-MAP2 (Abcam, ab5392, IF 1:500) rabbit anti-TOMM20 (11802-1-AP, Proteintech, IF 1:500), mouse anti-synaptophysin 1 (sc-17750, Santa Cruz, IF 1:500), chicken anti-neurofilament heavy polypeptide (ab4680, Abcam, IF 1:1,000), mouse anti-neurofilament marker (pan axonal, cocktail) (837904, BioLegend, IF 1:300), rabbit anti-chromogranin A (10529-1-AP, Proteintech, IF 1:500), mouse anti-Tau (Tau-1) (MAB3420, Millipore Sigma, IF 1:500). Secondary antibodies for IB and/or IF: HRP-conjugated goat anti-rabbit IgG (H+L) (111-035-003, Jackson Immuno Research, IB 1:5,000), Alexa Fluor 555-conjugated donkey anti-mouse IgG (A-31570, Thermo Scientific, IF 1:1,000), Alexa Fluor 555-conjugated goat anti-rabbit IgG (A-21428, Thermo Scientific, IF 1:1,000), Alexa Fluor 488-conjugated goat anti-rabbit IgG (A-11008, Thermo Scientific, IF 1:1,000), Alexa Fluor 647-conjugated goat anti chicken IgY, (A-21449 Thermo Scientific, IF 1:1,000). When possible, cell edges were outlined by staining of actin with Alexa FluorTM 647-conjugated phalloidin (A22287, Thermo Scientific, IF 1:500). Fluoromount G with DAPI (Electron Microscopy Sciences) was used for mounting and nuclear staining.

### Melanin quantification

One million MNT-1 cells (WT, *BLOC1S1*-KO, or *BLOC1S1*-KO rescued with *BLOC1S1* cDNA) were lifted and centrifuged at 600 *x g* for 5 min. For spectrophotometric measurement of melanin, the pellet was washed twice with 1x PBS and extracted in 200 μl of 1M NaOH containing 10% (v/v) DMSO at 80 °C with gentle agitation for 2 h. Melanin was measured by absorption spectroscopy at 405-nm wavelength using a DeNovix DS-11 spectrophotometer. For flow cytometry-based measurement of melanin, cell pellets were resuspended in 1 mL PBS, passed through a cell-strainer cap into 5-mL polystyrene round-bottom tubes (Falcon #352235), and kept on ice. Tubes were vortexed vigorously and a total of 100,000 events were acquired on an LSRFortessa Cell Analyze (BD Bioscience) flow cytometer. The side scatter (SSC) parameter, which quantifies light scattering from intracellular compartments, was measured and cell population data were analyzed using FlowJo Software version 10.6.1 FlowJo, LLC). Cell counts were plotted against side scatter area (SSC-A) and the geometric mean of the SSC-A parameter recorded. Previous literature described a quantitative linear relationship between melanin levels and SSC light scattering.^29^ We also observed that decreases in melanin content measured by absorption spectroscopy corresponded with decreases in SSC light scattering.

## Results

### Clinical phenotyping

We studied 11 individuals from seven unrelated families (FI-FVII) demonstrating a neurodevelopmental disorder associated with biallelic variants in *BLOC1S1.* These include four previously reported individuals from three families (FII-FIV)^24^ and seven additional individuals from four families (FI and FV-FVII) newly identified in this study (Figure 1A). All the affected individuals had disease onset in the first year of life, except for two more mildly affected subjects from FV (Table S2). The presentation across subjects was significant for developmental delay and later intellectual disability, hypotonia manifesting with delayed early motor milestones and profound impact on activities of daily living (ADLs), cortical visual impairment with bilateral optic nerve atrophy, failure to thrive, and, in some cases, refractory epilepsy (Table S2 and Supplemental notes). Disease severity ranged from early death (FIV:1 and FVII:1), profound spastic tetraparesis with epileptic encephalopathy (FI:1, FII:1, FII:2, FIII:1, FVI:1), to a milder phenotype with preserved ambulation and single word communication (FV:1 and FV:2). Two fetuses (FVII:2a and FVII2b) underwent pregnancy termination at 20 weeks and four days gestation due to prenatal findings consistent with the severe presentation of their older sibling (FVII:1). FI:1 was noted to have pale complexion and FVII:1 was reported to have hypopigmentation, but both individuals lacked features of oculocutaneous albinism. FIII:1 and FV:2 had ocular findings consistent with mild ocular albinism, but both had reportedly normal skin pigmentation. Most subjects in whom ophthalmologic examination was performed had optic atrophy. Several subjects had mild transaminase elevations attributed to anticonvulsant use. Otherwise, there were no findings characteristic of HPS such as bleeding diathesis, pulmonary fibrosis, granulomatous colitis, or immunodeficiency (Table S2).

At least single MRIs were available for review from FI:1, FII:1, FII:2, FIII:1, FV:1, and FVI:1 (Figure 1B, Supplemental information). FIV:1 and FVII:1 died in early infancy, with no prior MRI performed. Similarly, only prenatal ultrasounds were available for FVII:2a and FVII:2b.

Hypomyelination with or without superimposed sequela of deep cerebral white matter dysmyelination or demyelination, corpus callosum abnormalities (hypoplasia, hypogenesis/dysgenesis, or agenesis), and brain volume loss were typical brain MR features.

### Identification of *BLOC1S1* variants

Variants in *BLOC1S1* (NM_001487.4), located on chromosome 12q13.2, were identified in all 11 individuals by genome or exome sequencing. FI:1 was found to have compound heterozygous variants, including maternally inherited c.206A>C, p.His69Pro (H69P) and paternally inherited c.359G>A, p.Gly120Glu (G120E) missense variants (Table 1). All the other individuals had homozygous variants (Table 1), in agreement with reported consanguinity.

Some of these variants were missense [FII:1, FII:2, FV:1, and FV:2; c.284C>A p.Ala95Asp (A95D); FIII:1, c.185T>C; p.Leu62Pro (L62P); FVI:1, c.347T>C; p.Leu116Pro (L116P)] (Table 1). FIV:1 had an intronic variant, predicted to cause loss of the donor splice site from exon 2 to exon 3, resulting in a predicted exon skipping event and frameshift that would likely be subject to nonsense-mediated decay [c.218+3A>G; p.Glu49Valfs6 (E49*)] (Table 1). These variants were absent or found only in the heterozygous state in the gnomADv2.1 population allele-frequency database. They were predicted to be damaging across multiple *in silico* variant prediction tools including REVEL, CADD, MutationTaster, and SIFT, meeting PP3 criteria according to ACMG/AMP^30,31^ (Table S3). The amino-acid residues altered in these variants are all highly conserved through species (Figure 2B), indicating that changes to such residues could be detrimental for protein function. FVII:1, FVII:2a, and FVII:2b had a homozygous deletion including exons 2-4 of *BLOC1S1* and the first exon of *RDH5*. LoF variants in *RDH5* (OMIM:601617) are associated with a congenital form of night blindness with rod system impairment characterized by the presence of white-yellow retinal lesions.^32^ The premature death and neurodevelopmental phenotype of FVII:1, FVII:2a, and FVII:2b are therefore most consistent with the deletion of *BLOC1S1*.

Finally, we evaluated the gene-disease relationship (GDR) evidence following the Clinical Genome Resource (ClinGen) Clinical Validity Framework.^35^ Briefly, the strength of evidence for a GDR can be categorized as Definitive, Strong, Moderate, Limited, or No Known Disease Relationship. Only genes with evidence of Moderate or above should be included in clinical diagnostic testing.^36^ For *BLOC1S1*, we determined a Moderate level of evidence, supporting its association to autosomal recessive *BLOC1S1*-related hypomyelinating leukodystrophy.

### In silico analyses of *BLOC1S1* variants

The BLOC1S1 protein (NCBI Reference Sequence: NP_001478.2) is ubiquitously expressed in different cells and tissues (https://www.proteinatlas.org/ENSG00000135441-BLOC1S1). First described as a 125-amino-acid (AAB37682.1) translation product of cDNA isolated from a human fetal-brain cDNA library,^37^ the canonical human isoform (NP_001478.2) is now reported to be a 153-amino-acid protein. The 125- and 153-amino-acid proteins differ in their N-terminal residues and are suggested to represent products of alternative initiation at nearby methionine residues (M1 and M29, respectively), rather than alternative splicing products. Interestingly, orthologs from other species more strongly align with the shorter protein, lacking the longer N-terminal sequence. A number of *BLOC1S1* splice isoforms have also been reported based on identified transcript and peptide hits from large scale genomic and proteomic studies. All of these are predicted to be small proteins (< 100 amino acids), except for a 142-amino-acid isoform (KAI2566184.1) which encodes a unique C-terminus, compared to the canonical isoform.

As of this writing, high-resolution, three-dimensional structures of human BLOC1S1 and of the human BLOC-1 and BORC complexes have not been experimentally determined in humans. However, secondary structure predictions using JPred4 (http://www.compbio.dundee.ac.uk/jpred4)^38^ indicate that the canonical 153-amino-acid BLOC1S1 protein comprises a long α-helix (amino acids ∼26-144) preceded by a disordered sequence. When alternative initiation produces the shorter 125-amino-acid form of this protein, the N-terminal disordered sequence is lost. Furthermore, AlphaFold Multimer (Evans et al., 2021) predicts that both BLOC-1 and BORC are structured as an elongated core assembled from the α-helical regions of the eight subunits, with additional disordered regions extending from the core (Figure 2C-F),^19^ In these structures, BLOC1S1 is predicted to interact with all other subunits, except for SNAPIN and DTNBP1 in BLOC-1, and BORCS5 and SNAPIN in BORC (Figure S1A). The most extensive interactions of BLOC1S1 are predicted to be with BLOC1S6 (pallidin) in BLOC-1 and with KXD1 (BORCS4) in BORC (Figure S1B). The BORC structural prediction matches well with the recently published structure of BORC from *Caenorhabditis elegans*, particularly with the N-terminal region of BLOC1S1 positioned at the inter-subunit interface of the complex.^39^ The *BLOC1S1* variants identified in our cohort are located at highly conserved positions (Figure 2B) on the α-helical region of the protein (Figure 2C-F). The missense variants (FI, FII, FIII, FV, FVI) are predicted to affect interactions with different subunits of BLOC-1 and BORC (Table S4), potentially altering the local structure or function of the complexes. Alternatively, the frameshift variant (FIV [c.218+3A>G; p.Glu49Valfs6 (E49*)] is predicted to induce both splice donor and splice acceptor loss (score > 0.6) in all *BLOC1S1* transcript isoforms annotated to harbor this particular variant (see Table S5, scores highlighted in red). In particular, transcript isoforms (ENST00000548925 & ENST00000549147) highly expressed in developing brain encoding either the canonical protein (NP_001478.2) or the alternatively spliced 142-amino-acid isoform (KAI2566184.1), respectively, are affected by this variant. This variant would thus cause early truncation of the protein. Similarly, the 12q13.2 deletion in family VII encompasses the entirety of *BLOC1S1* and is therefore expected to result in complete LoF and a more severe phenotype as seen in individuals FVII:1, FVII:2a, and FVII:2b.

### Lysosome distribution and autophagic defects in *BLOC1S1*-KO HeLa cells

Next, we performed assays to assess the function of *BLOC1S1* in human cells. Because the clinical phenotype associated with *BLOC1S1* variants is more consistent with BORC deficiency (i.e., predominant neurological presentation)^19,24,25^ than BLOC-1 deficiency (i.e., pigmentation and prolonged bleeding),^40,41,42,43,44,45,46,47,48,49,50,51,52,53,54,55^ we initially focused on assays of BORC function. To this end, we used CRISPR-Cas9 to knock out *BLOC1S1* in human cervical adenocarcinoma HeLa cells. Immunoblot analysis of WT cells revealed two BLOC1S1 species (Figure 3A, arrowheads) that may correspond to the two alternative initiation products of the canonical isoform mentioned above. *BLOC1S1* KO resulted in complete loss of both species without altering the levels of the BLOC-1-specific subunit BLOC1S5 (muted), and the BORC-specific subunits BORCS5 (myrlysin) and BORCS7 (diaskedin) (Figure 3A). In contrast, KO of the BLOC-1-specific subunit BLOC1S6 (pallidin) decreased the levels of BLOC1S5 but not those of BORCS5 and BORCS7, while KO of BORCS5 decreased the levels of BORCS7 but not those of BLOC1S5 (Figure 3A). *BLOC1S1* KO thus appears to be less destabilizing than other subunits of both complexes in HeLa cells. The immunoblot analysis also revealed increased levels of the autophagy protein LC3B, particularly of the lipidated form LC3B-II (Figure 3A), in the *BLOC1S1*-KO cells, indicating that these cells have defective autophagy.

**Figure 3.**
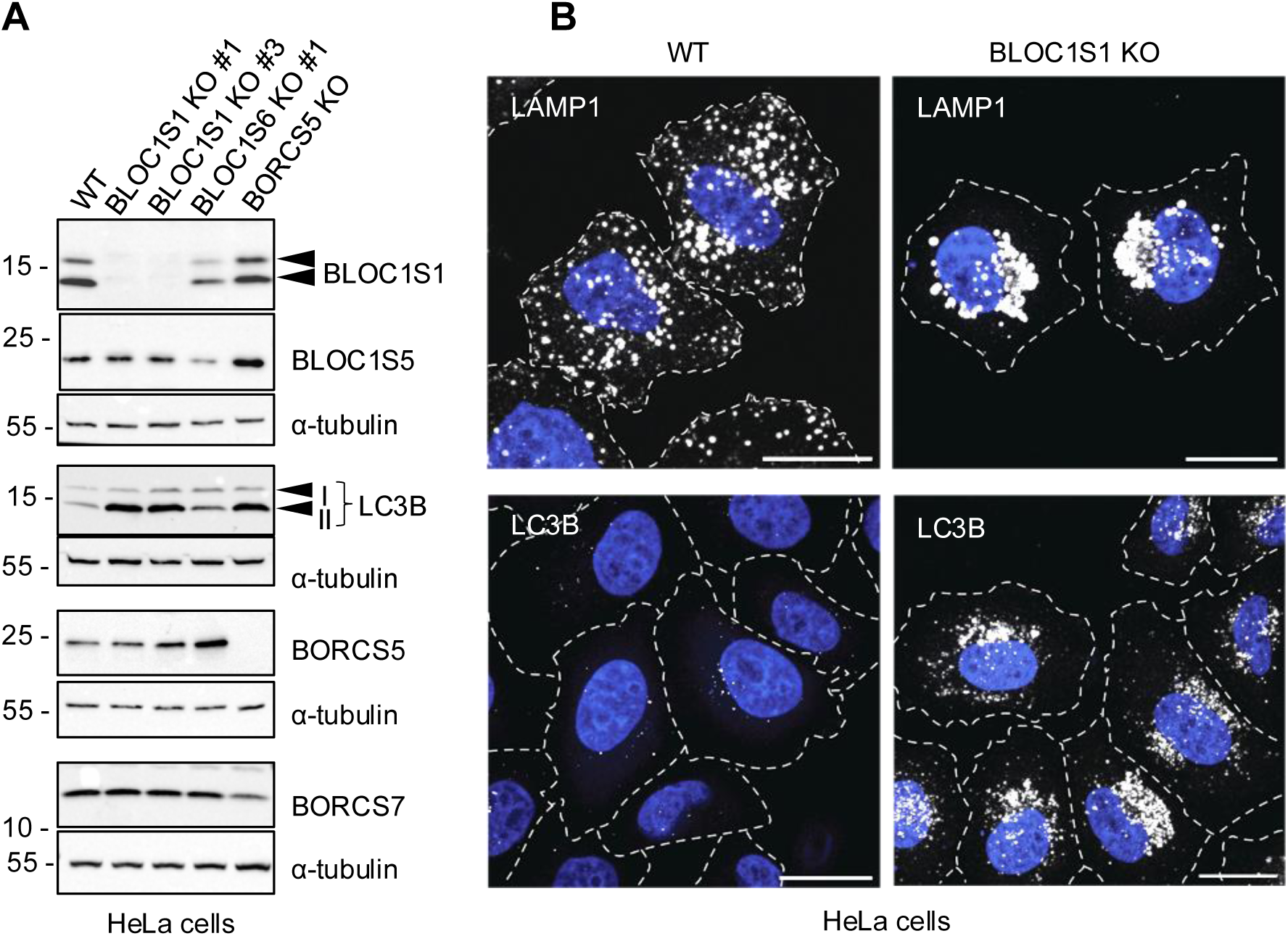
Characteristics of *BLOC1S1*-KO HeLa cells **(A)** SDS-PAGE and immunoblot analysis of WT and *BLOC1S1*-KO HeLa cells using antibodies to the endogenous proteins indicated on the right. α-tubulin was used as a control. Two isoforms of BLOC1S1 and the I and II forms of the autophagy protein LC3B are indicated by arrowheads on the right. The positions of molecular mass markers (in kDa) are indicated on the left. **(B)** WT and *BLOC1S1*-KO HeLa cells were fixed, permeabilized and immunostained for the endogenous lysosomal membrane protein LAMP1 and LC3B. Nuclei were labeled with DAPI (blue). Cell edges were outlined by staining of actin with fluorescent phalloidin or by background fluorescence (not shown, dashed lines). Images were obtained by confocal fluorescence microscopy. Scale bars: 20 μm.

Immunofluorescence microscopy for the endogenous lysosomal marker LAMP1 revealed that *BLOC1S1*-KO caused redistribution of lysosomes toward the perinuclear region of the cell (Figure 3B). This observation agrees with previous findings in cells lacking other subunits of BORC, and reflects the function of BORC in ARL8-dependent coupling of lysosomes to kinesins-1 and −3 for anterograde movement toward the peripheral cytoplasm.^4,16,19,21^ In line with the immunoblot analysis (Figure 3A), we also observed a large increase in staining for endogenous LC3B (Figure 3B). This phenotype is also similar to that of cells lacking other BORC subunits, and is indicative of the role of BORC in HOPS-dependent fusion of lysosomes with autophagosomes for eventual lysosomal degradation of LC3B.^19,21^

### Axonal alterations in *BLOC1S1* KO iPSC-derived neurons

Since the clinical phenotype related to *BLOC1S1* variants is mainly neurological, we next used CRISPR-Cas9 to KO *BLOC1S1* in human iPSCs for differentiation into neurons (i3Neurons). The human iPSC line used in these experiments expresses *NGN2* under the control of a doxycycline-inducible promoter, allowing for their differentiation into cortical-type, glutamatergic i3Neurons upon culture in doxycycline-containing medium.^27,28^ After 20 days in culture, immunoblot analysis showed that the BLOC1S1-KO i3Neurons expressed no BLOC1S1 and reduced levels of BLOC1S5, BORCS5 and BORCS7 (Figure 4A). This latter result contrasts with that in HeLa cells, in which the levels of BLOC1S5, BORCS5 and BORCS7 were not decreased upon *BLOC1S1* KO (Figure 3A). The subunits of both BLOC-1 and BORC are thus less stable in i3Neurons relative to HeLa cells when BLOC1S1 is absent.

**Figure 4.**
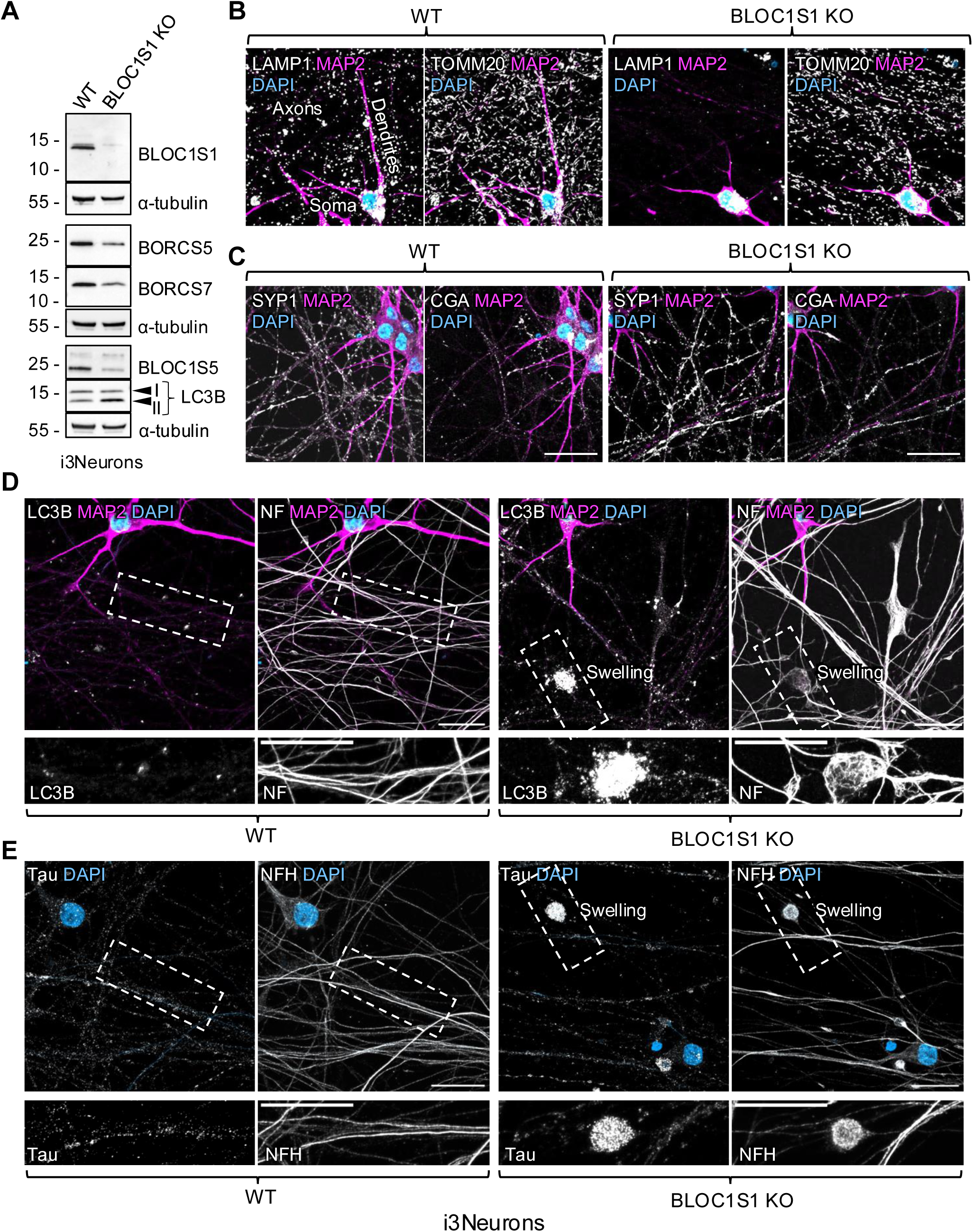
Characteristics of *BLOC1S1*-KO i3Neurons **(A)** SDS-PAGE and immunoblot analysis of WT and *BLOC1S1*-KO i3Neurons using antibodies to the endogenous proteins indicated on the right. α-tubulin was used as a control. Only one isoform of BLOC1S1 is observed in these neurons. The I and II forms of the autophagy protein LC3B are indicted by arrowheads on the right. The positions of molecular mass markers (in kDa) are indicated on the left. **(B, C)** WT and *BLOC1S1*-KO i3Neurons were cultured for 20 days on glass coverslips, fixed, permeabilized and immunostained for the endogenous somatodendritic marker MAP2 (magenta), lysosomal membrane protein LAMP1 (grayscale), and mitochondrial protein TOMM2 (grayscale) (B), or MAP2 (magenta), the synaptic vesicle protein SYP1 (grayscale), and the dense-core vesicle protein CGA (grayscale) (C). Nuclei were stained with DAPI (blue). Scale bars: 20 μm. The soma, dendrites and axons are indicated on the first image. **(D)** *Top row*: WT and *BLOC1S1*-KO i3Neurons were cultured for 20 days on glass coverslips, fixed, permeabilized and immunostained for endogenous MAP2 (magenta), LC3B (grayscale) and neurofilaments (NF) (grayscale). Nuclei were labeled with DAPI (blue). Scale bars: 20 μm. *Bottom row*: Enlarged images of the boxed areas in the upper row stained for the indicated markers. Scale bars: 20 μm. **(E)** *Top row*: WT and *BLOC1S1*-KO i3Neurons were cultured for 20 days on glass coverslips, fixed, permeabilized and immunostained with Tau-1 antibody (grayscale) and neurofilament heavy chain (NFH) antibody (grayscale). Scale bars: 20 μm. *Bottom row*: Enlarged images of the boxed areas in the upper row stained for the indicated markers. Scale bars: 20 μm. All images were obtained by confocal fluorescence microscopy.

Immunofluorescence microscopy for endogenous LAMP1 revealed a dramatic reduction in lysosomal vesicles within the axons of *BLOC1S1* KO compared to wild type (WT) i3Neurons (Figure 4B). In contrast, the absence of *BLOC1S1* did not impact the presence of mitochondria labeled for TOMM20 (Figure 4B), synaptic vesicles (SVs) labeled for synaptophysin 1 (SYP1), or dense core vesicles (DCVs) labeled for chromogranin A (CGA) in the axon (Figure 4C).

Moreover, the *BLOC1S1*-KO i3Neurons exhibited increased staining for LC3B in axons co-stained for neurofilament proteins (Figure 4D). LC3B accumulation was most pronounced within large axonal swellings, which exhibited abnormal organization of neurofilaments (Figure 4D) and intense staining for the microtubule associated protein and axonal degeneration marker tau (Figure 4E). These characteristics of *BLOC1S1*-KO i3Neurons resemble those of i3Neurons and rat hippocampal and cortical neurons lacking BORCS5, BORCS7 or BORCS8, underscoring the role of BORC in promoting transport of lysosomal vesicles into the axon for maintenance of axonal homeostasis.^17,18,19^

### Defective expression, assembly and/or BORC-related functions of *BLOC1S1* variants

Next, we examined the levels and assembly of six of the myc-tagged *BLOC1S1* variants (E49*, L62P, H69P, A95D, G120E, and A115S variant) expressed by transfection in human embryonic kidney HEK293T cells. Immunoblot analysis showed that all the missense variants were expressed at levels similar to those of WT *BLOC1S1*, while the frameshift variant E49* was expressed at much lower levels (Figure 5A, B, cell extracts). Immunoprecipitation using an antibody to the myc epitope followed by immunoblotting with antibodies to endogenous BORCS5, BORCS7 and BLOC1S5 allowed us to assess the assembly of the variants into the BORC and BLOC-1 complexes. We observed that whereas the H69P, A95D, and G120E variants assembled normally, the L62P and E49* variants exhibited reduced assembly with both BORC and BLOC-1 subunits (Figure 5A, B). The likely benign A115S variant also assembled normally.

**Figure 5.**
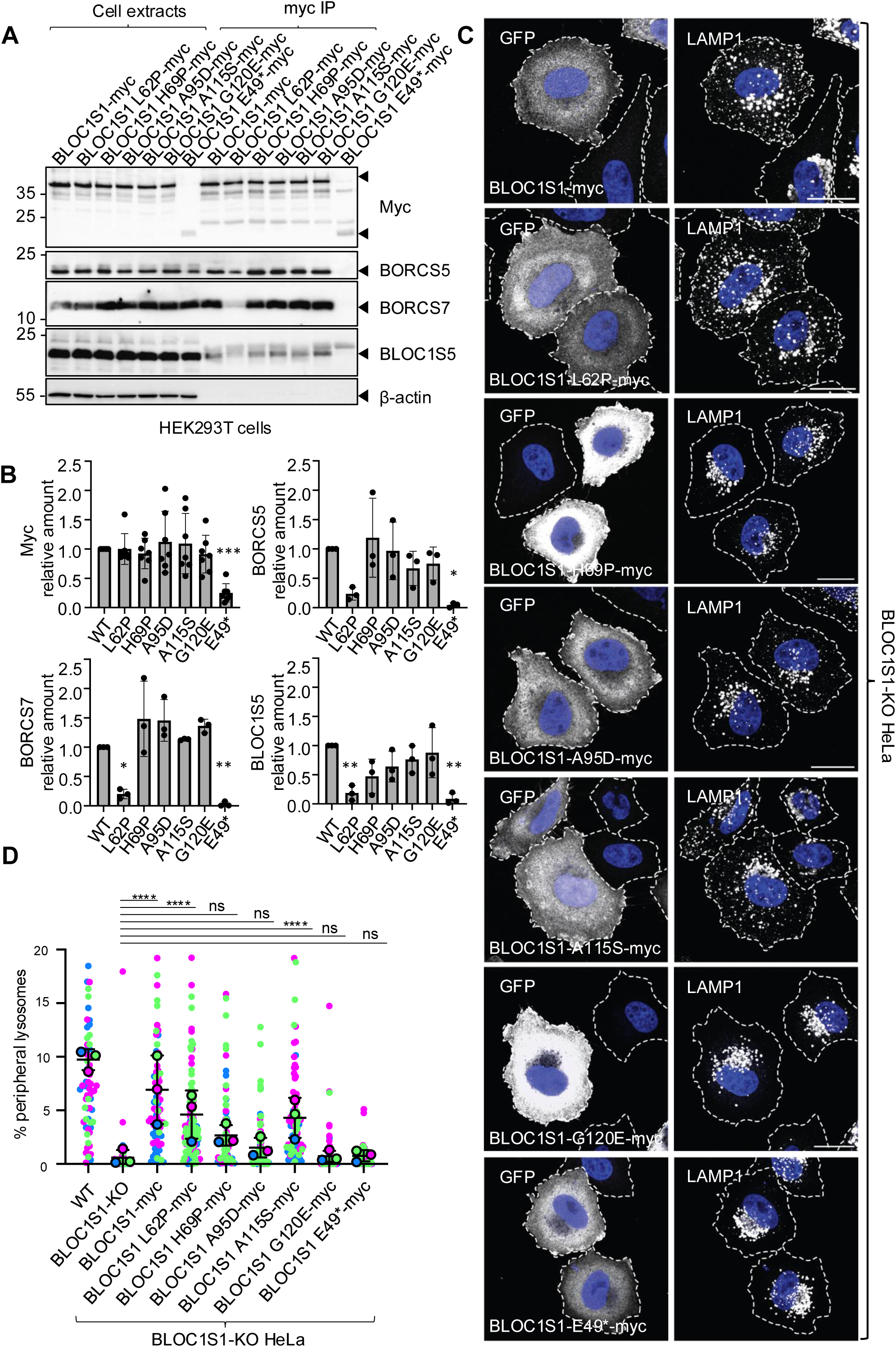
Assembly and lysosome-dispersal activity of *BLOC1S1* variants **(A)** HEK293T cells were transfected with plasmids encoding the indicated myc-tagged *BLOC1S1* variants and subjected to immunoprecipitation with antibody to the myc epitope. Cell extracts (10%) and immunoprecipitates (myc IP) were analyzed by SDS-PAGE and immunoblotting (IB) for the myc epitope and the endogenous proteins indicated on the right. β-actin was used as a control. Arrowheads point to the specific proteins. The positions of molecular mass markers (in kDa) are indicated on the left. **(B)** Quantification from three to seven independent experiments such as that shown in panel A. Values are the mean ± SD. Statistical significance was calculated by one-way ANOVA followed by multiple comparisons using Dunnett’s test. \**P* < 0.05; \*\**P* < 0.01; \*\*\**P* <0.001. **(C)** *BLOC1S1*-KO HeLa cells were co-transfected with plasmids encoding the indicated myc-tagged *BLOC1S1* variants and GFP (to identify transfected cells; grayscale). Cells were subsequently fixed, permeabilized and immunostained for the endogenous lysosomal membrane protein LAMP1. Nuclei were labeled with DAPI (blue). Cell edges were outlined by staining of actin with fluorescent phalloidin (not shown, dashed lines). Images were obtained by confocal fluorescence microscopy. Scale bars: 20 μm. **(D)** Quantification by shell analysis^69^ of peripheral LAMP1 signal in *BLOC1S1*-KO HeLa cells expressing different *BLOC1S1* variants from three experiments such as that shown in panel C. Data are represented as SuperPlots^70^ showing the individual data points, the mean from each experiment, and the mean ± SD of the means. Statistical significance was calculated by one-way ANOVA followed by multiple comparisons using Tukey’s test. \*\*\*\**P* < 0.0001; ns: not significant.

To assess the activity of the E49*, L62P, H69P, A95D, and G120E variants, we transfected *BLOC1S1*-KO HeLa cells with plasmids encoding the different myc-tagged variants and examined the distribution of LAMP1 in the transfected cells. These experiments showed that while expression of WT, L62P or A115S *BLOC1S1* rescued the peripheral distribution of lysosomes, expression of H69P, A95D, G120D and E49* *BLOC1S1* did not (Figure 5C, D). In addition, we examined the ability of the myc-tagged *BLOC1S1* variants to restore low basal levels of LC3B upon expression in *BLOC1S1*-KO HeLa cells. We found that expression of WT, L62P, H69P, and A95D *BLOC1S1* rescued LC3B levels, while expression of G120D and E49* resulted in partial or no rescue (Figure 6A, B).

**Figure 6.**
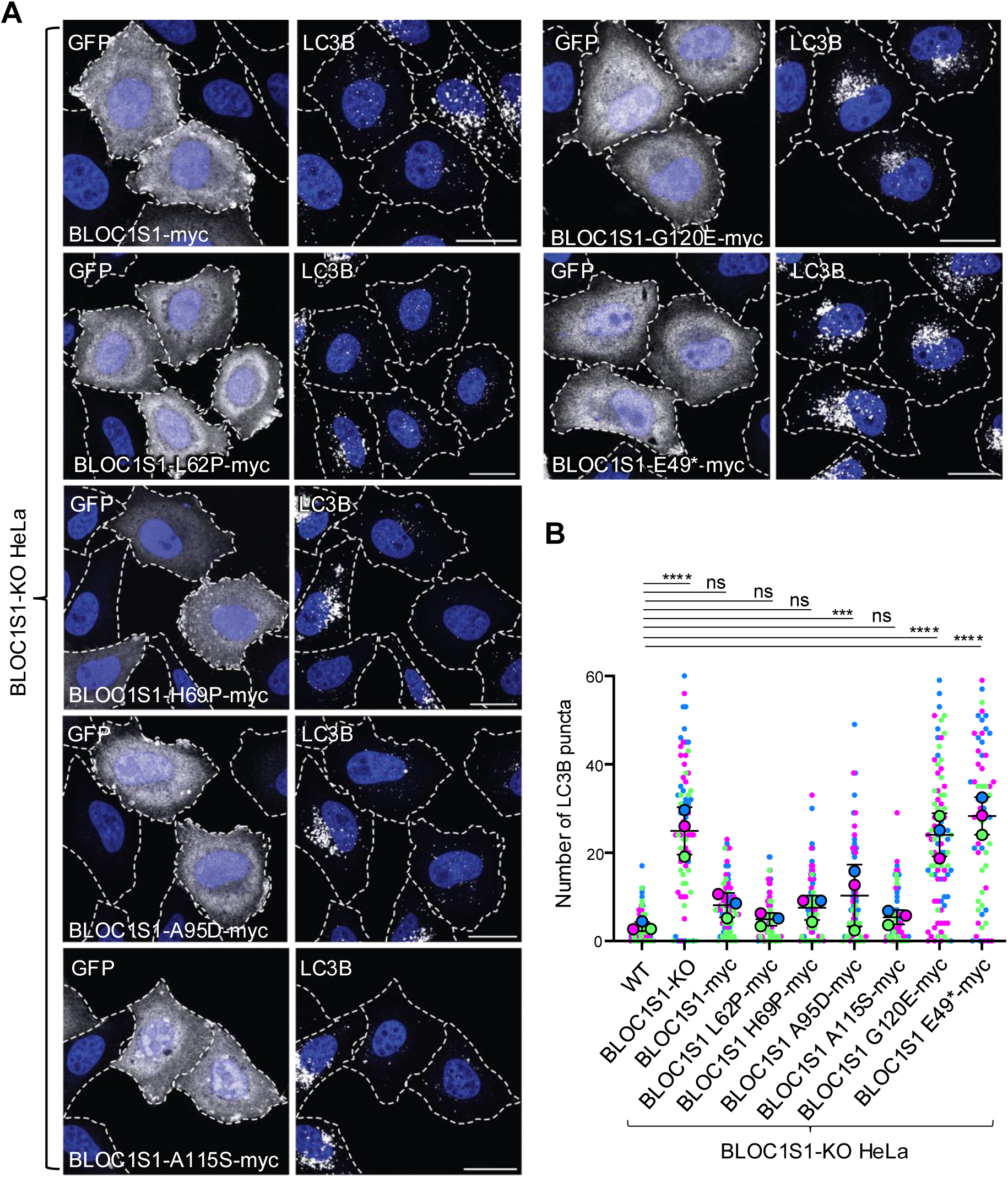
Autophagy-promoting activity of *BLOC1S1* variants **(A)** *BLOC1S1*-KO HeLa cells were co-transfected with plasmids encoding the indicated myc-tagged *BLOC1S1* variants and GFP (to identify transfected cells; grayscale). Cells were subsequently fixed, permeabilized and immunostained for the endogenous autophagy protein LC3B. Nuclei were labeled with DAPI (blue). Cell edges were outlined based on background fluorescence (not shown, dashed lines). Images were obtained by confocal fluorescence microscopy. Scale bars: 20 μm. **(B)** Quantification of LC3B puncta per cell in *BLOC1S1*-KO HeLa cells expressing *BLOC1S1* variants from three experiments such as that shown in panel A. Data were represented as SuperPlots^70^ showing the individual data points, the mean from each experiment, and the mean ± SD of the means. Statistical significance was calculated by one-way ANOVA followed by multiple comparisons using Tukey’s test. \*\*\**P* < 0.001; \*\*\*\**P* < 0.0001; ns: not significant.

Taken together, these results demonstrated that most of the *BLOC1S1* variants tested exhibit some kind of BORC-related impairment (Table 2). The E49* frameshift variant is the most defective, showing reduced expression, assembly and ability to rescue normal lysosome distribution and LC3B levels. Except for A115S, a likely benign variant that behaves indistinguishably from WT in all the assays, the other missense variants displayed various combinations of defective expression, assembly, lysosome dispersal and/or autophagy. These findings provide PS3-level evidence in favor of the pathogenicity of all *BLOC1S1* variants characterized here (Table 3).

**Table 2.** Summary of *BLOC1S1* variant cellular phenotypes.

| Subjects | FI:1 | FI:1 | FII:1, FII:2,<br>FV:1, FV:2 | FIII:1 | FIV:1 | Control |
| --- | --- | --- | --- | --- | --- | --- |
| Amino-acid changes | p.His69Pro (H69P) | p.Gly120Glu (G120E) | p.Ala95Asp (A95D) | p.Leu62Pro (L62P) | p.Glu49Valfs6 (VWPRPT Stop) (E49*) | p.Ala115Ser (A115S) |
| Expression | Normal | Normal | Normal | Normal | Reduced | Normal |
| Assembly | Normal | Normal | Normal | Reduced | Absent | Normal |
| Rescue of lysosome dispersal | Absent | Absent | Absent | Normal | Absent | Normal |
| Rescue of LC3B levels | Normal | Absent | Reduced | Normal | Absent | Normal |
| Rescue of pigmentation | Normal | Reduced | Normal | Normal | Reduced | Normal |

**Table 3.** Summary of *BLOC1S1* variant classification.

| BLOC1S1 variant | Classification | Criteria (per ACMG/AMP/ClinGen SVI guidelines) |
| --- | --- | --- |
| c.206A>C p.His69Pro | VUS | PM2_Supp, PS3_Supp |
| c.359G>A p.Gly120Glu | VUS | PM2_Supp, PP3_Mod, PS3_Supp |
| c.284C>A p.Ala95Asp | Likely Pathogenic | PM2_Supp, PM3_Mod, PP1_Mod, PS3_Supp |
| c.185T>C p.Leu62Pro | VUS | PM2_Supp, PP3_Mod, PM3_Supp, PS3_Supp |
| c.218+3A>G p.? | VUS | PM2_Supp, PP3_Supp, PM3_Supp, PS3_Mod |
| c.347T>C p.Leu116Pro | Likely Pathogenic | PM2_Supp, PP3_Str, PM3_Supp |
| c.343G>T p.Ala115Ser | Likely Benign | BP4_Supp, BS3_Supp, PM2_Supp |
| 3.9kb 12q13.2 deletion | VUS | 4F (per PMID: 31690835) |

### Effects of *BLOC1S1* KO or variants in a melanocytic cell line

Whereas KO or LoF variants in *BLOC1S1* orthologs in mouse,^56^ Drosophila,^57^ and zebrafish^58^ cause defective pigmentation, this has not been demonstrated in humans. A pale skin relative to family members (FI:1), hypopigmentation (FVII:1), and ocular findings consistent with ocular albinism (FIII:1 and FV:2) were observed in some of the individuals harboring *BLOC1S1* variants reported here, but these pigmentation defects are milder than those in HPS. To analyze the role of BLOC1S1 in human pigmentation, we used CRISPR-Cas9 to KO *BLOC1S1* in MNT-1 cells (Figure 7A), a human melanocytic cell line commonly used in studies of pigmentation.^59^ We observed that *BLOC1S1*-KO MNT-1 cells had no detectable BLOC1S1 protein (Figure 7A). These cells exhibited reduced melanin content relative to WT MNT-1 cells, as assessed by the lighter color of cell pellets (Figure 7B), decreased side-scatter (SSC) by flow cytometry (Figure 7C and D),^29^ and decreased absorbance of cell extracts at 405 nm detected by spectrophotometry (Figure 7E, two leftmost columns). Since the spectrophotometry assay was more sensitive to detect differences between *BLOC1S1*-KO and WT cells, we used it to assess the activity of the E49*, L62P, H69P, A95D, and G120E variants. This was done by infecting *BLOC1S1*-KO MNT-1 cells with viruses encoding the different variants, and measuring melanin content. The results showed that expression of all *BLOC1S1* variants rescued melanin content in *BLOC1S1*-KO cells, completely in the case of L62P, H69P, A95D or A115S *BLOC1S1* and partially in the case of G120D and E49* *BLOC1S1* (Figure 7E). Therefore, most of the variants are more defective in BORC-related functions (Figures 5 and 6) than BLOC-1-related functions (Figure 7), consistent with the clinical presentation of the *BLOC1S1* variants as a neurological disorder with little or no evidence of reduced pigmentation.

**Figure 7.**
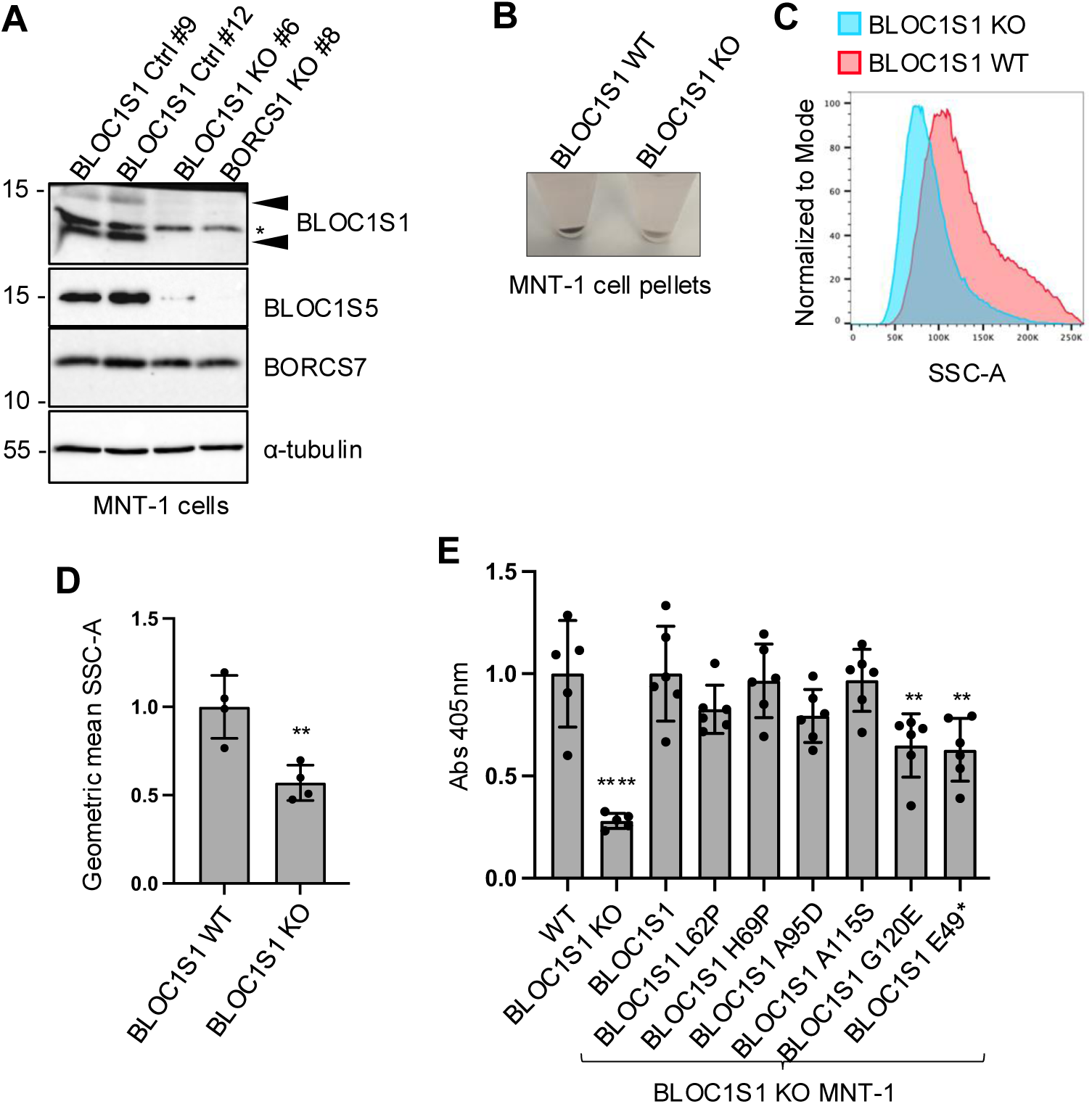
Pigmentation activity of *BLOC1S1* variants **(A)** SDS-PAGE and immunoblot analysis of WT and *BLOC1S1*-KO MNT-1 cells using antibodies to the endogenous proteins indicated on the right. α-tubulin was used as a control. Two variants of *BLOC1S1* are indicated by arrowheads. The positions of molecular mass markers (in kDa) are indicated on the left. **(B)** WT and *BLOC1S1*-KO MNT-1 cell pellets. Notice the lighter pigmentation of the KO cells. **(C)** Flow cytometry of WT and *BLOC1S1*-KO MNT-1 cells. The x-axis indicates the number of cells normalized to the mode (or peak) of the graph. The y-axis indicates side scatter area (SSC-A). **(D)** Bar graph representing the geometric mean ± SD values of the SSC-A parameter from three independent flow cytometry experiments like that shown in panel C. **(E)** Bar graph representing the light absorbance at 405 nm measured after melanin extraction. *BLOC1S1*-KO MNT-1 cells were rescued with lentiviruses carrying the indicated protein. WT and *BLOC1S1*-KO MNT-1 cells were used as controls. Data from five or six independent experiments are represented as the mean ± SD. Statistical significance was calculated by one-way ANOVA followed by multiple comparisons using Tukey’s test. \*\**P* < 0.005; \*\*\*\**P* < 0.0001; ns: not significant.

## Discussion

Herein, we present 11 individuals from seven unrelated families, including new phenotypics details for four previously reported individuals.^24^ All individuals are affected by a severe neurodevelopmental disorder and harbor biallelic variants of *BLOC1S1*. The disorder presents with early infantile onset and is characterized by deficient myelination, global developmental delay, intellectual disability, hypotonia, epilepsy, and visual impairment. The severity ranged from early death to a milder form with preserved ambulation and single-word communication. These neurological features are consistent with those recently described in individuals with pathogenic variants in *BORCS8.*^19^ Given that both *BLOC1S1* and *BORCS8* encode subunits of the hetero-octameric BORC complex,^4^ these findings begin to define a broader clinical entity characterized by BORC deficiency.

The visual impairment in individuals harboring *BLOC1S1* variants included bilateral optic atrophy in most individuals, and features of ocular albinism in two individuals (FIII:1 and FV:2). In addition, one individual had a pale complexion (FI:1) and one had hypopigmentation (FVII:1). The ocular albinism and pale complexion are reminiscent of those in HPS types 7, 8, 9 and 11 caused by variants in the *DTNBP1*, *BLOC1S3*, *BLOC1S6* and *BLOC1S5* genes, respectively.^41,40,42,43,44,45,46,47,48,49,50,54,51,55,52,53^ All these genes encode subunits of the related hetero-octameric complex BLOC-1.^1,2,3^ These findings are consistent with BLOC1S1 being a subunit of BLOC-1^1,2,3^ in addition to BORC.^4^ However, the pigmentation defects in the individuals harboring *BLOC1S1* variants were notably milder than those in HPS, and other HPS features such as bleeding diathesis were not observed. These findings suggest that the *BLOC1S1* variants reported here have a more significant impact on BORC compared to BLOC-1. Thus, individuals with BLOC1S1 variants exhibit clinical and functional phenotypes more greatly impacted by BORC than BLOC-1 deficits.

Three of the individuals from one family (FVII) described here had complete deletion of *BLOC1S1* and died on the first day after birth (FVII:1) or underwent pregnancy termination at 20 weeks and four days (FVII:2a and FVII:2b). However, these individuals also had complete deletions of three other neighboring genes, making it uncertain if their premature death was solely due to the deletion of *BLOC1S1*. In any event, this suggests that all the other *BLOC1S1* variants studied here are hypomorphic. The residual activity of these variants may be sufficient for the function of BLOC-1 in the biogenesis of LROs like melanosomes and platelet dense bodies, but insufficient for the function of BORC in the development of the central nervous system (CNS).

Brain or neuronal defects have been reported in mice,^56^ Drosophila^57^ and *C. elegans,*^60^ and defective pigmentation has been observed in mice,^56^ Drosophila^57^ and zebrafish,^58^ with KO or variants in *BLOC1S1* orthologs. These findings indicate that the role of *BLOC1S1* in neurodevelopment and pigmentation is broadly conserved across metazoans. However, the effects of *BLOC1S1* variants had not been previously described in human cells. Our experiments demonstrate that KO of *BLOC1S1* in human HeLa cells leads to redistribution of lysosomes towards the perinuclear cytoplasm and increased levels of the autophagy marker LC3-II. These phenotypes are characteristic of defective BORC/ARL8-dependent coupling of lysosomes to kinesins 1 and 3^4,16^ and to the HOPS complex,^20,21^ respectively. Moreover, we found that *BLOC1S1* KO in human iPSC-derived neurons reduced the transport of lysosomal vesicles into the axon, also consistent with the role of BORC and ARL8 in mediating kinesin-driven transport of these organelles into the axon.^18,19,61^ Additionally, we observed increased levels of neuronal LC3-II, with particularly pronounced accumulation in aberrant swellings along the axon shaft. These swellings were filled with the microtubule protein tau and the neurofilament protein NFH, all characteristics of axonal dystrophy or degeneration.^62,63^ These neuronal defects may thus contribute to the neurodevelopmental aspects of the disease.

While *BLOC1S1* KO in human cells served as a model for the *BLOC1S1*-deletion in FVII:1, FVII:2 and FVII:3 individuals, the rescue of peripheral lysosome distribution and basal LC3-II levels in HeLa cells was used to assess the properties of several frameshift or missense variants. We found that all the tested variants exhibited varying defects in expression, assembly, lysosome dispersal and/or autophagy. Among them, the frameshift variant E49* showed the most severe deficiencies, with little to no activity in all assays. The missense variants demonstrated defects in one or more of the assessed parameters, including expression, assembly and/or function. These findings provide relevant evidence in favor of the pathogenicity of all the tested variants.^30,31^

*BLOC1S1* KO in the human melanocytic cell line MNT-1 also resulted in reduced pigmentation. The observation that the individuals harboring *BLOC1S1* variants had only mild or no pigmentation defects supports the conclusion that their *BLOC1S1* variants retain sufficient activity to maintain melanosome development, albeit with some reduction. This is consistent with the ability of all identified *BLOC1S1* variants to completely or partially rescue the reduced pigmentation of *BLOC1S1*-KO MNT-1 cells. Mice with heart- or liver-specific KO of *Bloc1s1* exhibit defects in the corresponding organs.^64,65,66,67,68^ However, individuals harboring *BLOC1S1* variants exhibited no obvious cardiac or hepatic defects, suggesting that the *BLOC1S1* variants retain sufficient activity in these organs as well. Predominant neurological presentation in *BLOC1S1*-associated human disease may reflect the greater vulnerability of neurons and possibly glial cells to disruptions in intracellular protein and organelle trafficking.

In conclusion, we expand BORC-associated disease through the description of 11 subjects with *BLOC1S1* relevant biallelic variants. Individuals harboring *BLOC1S1* variants have a primarily neurologic phenotype of early infantile onset developmental delay, with intellectual and motor impairments. Clinical course is complicated by epilepsy and early mortality. In addition, individuals harboring *BLOC1S1* variants have profound CNS hypomyelination. A subset of individuals also show features of mild oculocutaneous albinism, which is explained by the dual role of the BLOC1S1 subunit in BLOC-1 and BORC. Altogether, our clinical and functional studies establish BLOC1S1 bi-allelic variants as a new genetic cause of a lysosomal disorder characterized by hypomyelinating leukodystrophy with epileptic encephalopathy.

## Data Availability

All data are available in the main text or Supplementary Information. Reagents generated in this study are available upon reasonable request. Further information and requests for resources and reagents should be directed to the corresponding author. Source data are provided with this article.

## Acknowledgements

The Vanderver lab was supported by funding from the NINDS and NCATS (U54NS115052). The Bonifacino lab was supported by the Intramural Program of the NICHD (ZIA HD001607). The Barakat lab was supported by the Netherlands Organisation for Scientific Research (ZonMw Vidi, grant 09150172110002). The Bhoj and Ahrens-Nicklas labs were supported by a Chan Zuckerberg Initiative Neurodegeneration Challenge Collaborative Pairs grant. The Simons lab was supported by the Medical Research Future Fund (ARG76368). JLB was supported by NIH (U54NS115052). We thank Stichting 12q (https://stichting12q.nl/) for supporting our work related to disorders on chromosome 12q. The funders had no role in study design, data collection and analysis, decision to publish or preparation of the manuscript.

## Author contributions

RDP, CDG, AV and JSB conceived the study with input from all authors. CDG, RJT, CS, RM, EZ, AR, KT, AMBA and AV performed variant identification and clinical genotyping. GH, AP, BD, JLS, KM, JLB, RJT, NAS, PD, MBM, HB, HKJ, AHI, HH, HO, HB, MM, MA, SS, EZ, TSB, AV, EB, RAN, EZ, AR, KT, AMBA, and RM analyzed clinical data. AV and MW interpreted and described neuroimaging. RDP and NHS carried out all the cellular experiments. CDW and ET performed structural predictions. RDP, CDG, AV and JSB wrote the manuscript with input from all the authors. All authors read and edited the manuscript.

## Supplemental information

Supplemental information includes Supplemental note: case reports, Figure S1, and Tables S1-S5.

## References

1. Falcón-Pérez, J.M., Starcevic, M., Gautam, R., and Dell’Angelica, E.C. (2002). BLOC-1, a novel complex containing the pallidin and muted proteins involved in the biogenesis of melanosomes and platelet-dense granules. J Biol Chem 277, 28191–28199.

2. Moriyama, K., and Bonifacino, J.S. (2002). Pallidin is a component of a multi-protein complex involved in the biogenesis of lysosome-related organelles. Traffic 3, 666–677.

3. Starcevic, M., and Dell’Angelica, E.C. (2004). Identification of snapin and three novel proteins (BLOS1, BLOS2, and BLOS3/reduced pigmentation) as subunits of biogenesis of lysosome-related organelles complex-1 (BLOC-1). J Biol Chem 279, 28393–28401.

4. Pu, J., Schindler, C., Jia, R., Jarnik, M., Backlund, P., and Bonifacino, J.S. (2015). BORC, a multisubunit complex that regulates lysosome positioning. Dev Cell 33, 176–188.

5. Hartwig, C., Monis, W.J., Chen, X., Dickman, D.K., Pazour, G.J., and Faundez, V. (2018). Neurodevelopmental disease mechanisms, primary cilia, and endosomes converge on the BLOC-1 and BORC complexes. Dev Neurobiol 78, 311–330.

6. Dell’Angelica, E.C., Mullins, C., Caplan, S., and Bonifacino, J.S. (2000). Lysosome-related organelles. FASEB J 14, 1265–1278.

7. Marks, M.S., Heijnen, H.F., and Raposo, G. (2013). Lysosome-related organelles: unusual compartments become mainstream. Curr Opin Cell Biol 25, 495–505.

8. Huang, L., Kuo, Y.M., and Gitschier, J. (1999). The pallid gene encodes a novel, syntaxin 13-interacting protein involved in platelet storage pool deficiency. Nat Genet 23, 329–332.

9. Zhang, Q., Li, W., Novak, E.K., Karim, A., Mishra, V.S., Kingsmore, S.F., Roe, B.A., Suzuki, T., and Swank, R.T. (2002). The gene for the muted (mu) mouse, a model for Hermansky-Pudlak syndrome, defines a novel protein which regulates vesicle trafficking. Hum Mol Genet 11, 697–706.

10. Ciciotte, S.L., Gwynn, B., Moriyama, K., Huizing, M., Gahl, W.A., Bonifacino, J.S., and Peters, L.L. (2003). Cappuccino, a mouse model of Hermansky-Pudlak syndrome, encodes a novel protein that is part of the pallidin-muted complex (BLOC-1). Blood 101, 4402–4407.

11. Gwynn, B., Martina, J.A., Bonifacino, J.S., Sviderskaya, E.V., Lamoreux, M.L., Bennett, D.C., Moriyama, K., Huizing, M., Helip-Wooley, A., Gahl, W.A., et al. (2004). Reduced pigmentation (rp), a mouse model of Hermansky-Pudlak syndrome, encodes a novel component of the BLOC-1 complex. Blood 104, 3181–3189.

12. Di Pietro, S.M., Falcón-Pérez, J.M., Tenza, D., Setty, S.R., Marks, M.S., Raposo, G., and Dell’Angelica, E.C. (2006). BLOC-1 interacts with BLOC-2 and the AP-3 complex to facilitate protein trafficking on endosomes. Mol Biol Cell 17, 4027–4038.

13. Delevoye, C., Heiligenstein, X., Ripoll, L., Gilles-Marsens, F., Dennis, M.K., Linares, R.A., Derman, L., Gokhale, A., Morel, E., Faundez, V., et al. (2016). BLOC-1 Brings Together the Actin and Microtubule Cytoskeletons to Generate Recycling Endosomes. Curr Biol 26, 1–13.

14. Jani, R.A., Di Cicco, A., Keren-Kaplan, T., Vale-Costa, S., Hamaoui, D., Hurbain, I., Tsai, F.C., Di Marco, M., Macé, A.S., Zhu, Y., et al. (2022). PI4P and BLOC-1 remodel endosomal membranes into tubules. J Cell Biol 221, e202110132.

15. Zhu, Y., Li, S., Jaume, A., Jani, R.A., Delevoye, C., Raposo, G., and Marks, M.S. (2022). Type II phosphatidylinositol 4-kinases function sequentially in cargo delivery from early endosomes to melanosomes. J Cell Biol 221, e202110114.

16. Guardia, C.M., Farías, G.G., Jia, R., Pu, J., and Bonifacino, J.S. (2016). BORC functions upstream of kinesins 1 and 3 to coordinate regional movement of lysosomes along different microtubule Tracks. Cell Rep 17, 1950–1961.

17. Snouwaert, J.N., Church, R.J., Jania, L., Nguyen, M., Wheeler, M.L., Saintsing, A., Mieczkowski, P., Manuel de Villena, F.P., Armao, D., Moy, S.S., et al. (2018). A mutation in the Borcs7 subunit of the lysosome regulatory BORC complex results in motor deficits and dystrophic axonopathy in mice. Cell Rep 24, 1254–1265.

18. De Pace, R., Britt, D.J., Mercurio, J., Foster, A.M., Djavaherian, L., Hoffmann, V., Abebe, D., and Bonifacino, J.S. (2020). Synaptic vesicle precursors and lysosomes are transported by different mechanisms in the axon of mammalian neurons. Cell Rep 31, 107775.

19. De Pace, R., Maroofian, R., Paimboeuf, A., Zamani, M., Zaki, M.S., Sadeghian, S., Azizimalamiri, R., Galehdari, H., Zeighami, J., Williamson, C.D., et al. (2024). Biallelic BORCS8 variants cause an infantile-onset neurodegenerative disorder with altered lysosome dynamics. Brain 147, 1751–1767.

20. Khatter, D., Raina, V.B., Dwivedi, D., Sindhwani, A., Bahl, S., and Sharma, M. (2015). The small GTPase Arl8b regulates assembly of the mammalian HOPS complex on lysosomes. J Cell Sci 128, 1746–1761.

21. Jia, R., Guardia, C.M., Pu, J., Chen, Y., and Bonifacino, J.S. (2017). BORC coordinates encounter and fusion of lysosomes with autophagosomes. Autophagy 13, 1648–1663.

22. Schleinitz, A., Pöttgen, L.A., Keren-Kaplan, T., Pu, J., Saftig, P., Bonifacino, J.S., Haas, A., and Jeschke, A. (2023). Consecutive functions of small GTPases guide HOPS-mediated tethering of late endosomes and lysosomes. Cell Rep 42, 111969.

23. Di Pietro, S.M., and Dell’Angelica, E.C. (2005). The cell biology of Hermansky-Pudlak syndrome: recent advances. Traffic 6, 525–533.

24. Bertoli-Avella, A.M., Kandaswamy, K.K., Khan, S., Ordonez-Herrera, N., Tripolszki, K., Beetz, C., Rocha, M.E., Urzi, A., Hotakainen, R., Leubauer, A., et al. (2021). Combining exome/genome sequencing with data repository analysis reveals novel gene-disease associations for a wide range of genetic disorders. Genet Med 23, 1551–1568.

25. Charng, W.L., Karaca, E., Coban Akdemir, Z., Gambin, T., Atik, M.M., Gu, S., Posey, J.E., Jhangiani, S.N., Muzny, D.M., Doddapaneni, H., et al. (2016). Exome sequencing in mostly consanguineous Arab families with neurologic disease provides a high potential molecular diagnosis rate. BMC Med Genomics 9, 42.

26. Vanderver, A., Bernard, G., Helman, G., Sherbini, O., Boeck, R., Cohn, J., Collins, A., Demarest, S., Dobbins, K., Emrick, L., et al. (2020). Randomized Clinical Trial of First-Line Genome Sequencing in Pediatric White Matter Disorders. Ann Neurol 88, 264–273.

27. Wang, C., Ward, M.E., Chen, R., Liu, K., Tracy, T.E., Chen, X., Xie, M., Sohn, P.D., Ludwig, C., Meyer-Franke, A., et al. (2017). Scalable Production of iPSC-Derived Human Neurons to Identify Tau-Lowering Compounds by High-Content Screening. Stem Cell Reports 9, 1221–1233.

28. Fernandopulle, M.S., Prestil, R., Grunseich, C., Wang, C., Gan, L., and Ward, M.E. (2018). Transcription Factor-Mediated Differentiation of Human iPSCs into Neurons. Curr Protoc Cell Biol 79, e51.

29. Bajpai, V.K., Swigut, T., Mohammed, J., Naqvi, S., Arreola, M., Tycko, J., Kim, T.C., Pritchard, J.K., Bassik, M.C., and Wysocka, J. (2023). A genome-wide genetic screen uncovers determinants of human pigmentation. Science 381, eade6289.

30. Richards, S., Aziz, N., Bale, S., Bick, D., Das, S., Gastier-Foster, J., Grody, W.W., Hegde, M., Lyon, E., Spector, E., et al. (2015). Standards and guidelines for the interpretation of sequence variants: a joint consensus recommendation of the American College of Medical Genetics and Genomics and the Association for Molecular Pathology. Genet Med 17, 405–424.

31. Brnich, S.E., Abou Tayoun, A.N., Couch, F.J., Cutting, G.R., Greenblatt, M.S., Heinen, C.D., Kanavy, D.M., Luo, X., McNulty, S.M., Starita, L.M., et al. (2019). Recommendations for application of the functional evidence PS3/BS3 criterion using the ACMG/AMP sequence variant interpretation framework. Genome Med 12, 3.

32. Yamamoto, H., Simon, A., Eriksson, U., Harris, E., Berson, E.L., and Dryja, T.P. (1999). Mutations in the gene encoding 11-cis retinol dehydrogenase cause delayed dark adaptation and fundus albipunctatus. Nat Genet 22, 188–191.

33. Hayashi, Y.K., Chou, F.L., Engvall, E., Ogawa, M., Matsuda, C., Hirabayashi, S., Yokochi, K., Ziober, B.L., Kramer, R.H., Kaufman, S.J., et al. (1998). Mutations in the integrin alpha7 gene cause congenital myopathy. Nat Genet 19, 94–97.

34. Schröder, J., Lüllmann-Rauch, R., Himmerkus, N., Pleines, I., Nieswandt, B., Orinska, Z., Koch-Nolte, F., Schröder, B., Bleich, M., and Saftig, P. (2009). Deficiency of the tetraspanin CD63 associated with kidney pathology but normal lysosomal function. Mol Cell Biol 29, 1083–1094.

35. Strande, N.T., Riggs, E.R., Buchanan, A.H., Ceyhan-Birsoy, O., DiStefano, M., Dwight, S.S., Goldstein, J., Ghosh, R., Seifert, B.A., Sneddon, T.P., et al. (2017). Evaluating the Clinical Validity of Gene-Disease Associations: An Evidence-Based Framework Developed by the Clinical Genome Resource. Am J Hum Genet 100, 895–906.

36. Bean, L.J.H., Funke, B., Carlston, C.M., Gannon, J.L., Kantarci, S., Krock, B.L., Zhang, S., Bayrak-Toydemir, P., Assurance, A.C.M.G.L.Q., and Committee (2020). Diagnostic gene sequencing panels: from design to report-a technical standard of the American College of Medical Genetics and Genomics (ACMG). Genet Med 22, 453–461.

37. Watanabe, T.K., Fujiwara, T., Shinomiya, H., Kuga, Y., Hishigaki, H., Nakamura, Y., and Hirai, Y. (1995). Molecular cloning of a novel human cDNA, RT14, containing a putative ORF highly conserved between human, fruit fly, and nematode. DNA Res 2, 235–237.

38. Drozdetskiy, A., Cole, C., Procter, J., and Barton, G.J. (2015). JPred4: a protein secondary structure prediction server. Nucleic Acids Res 43, W389–94.

39. Ge, X., Ren, J., Gu, K., Gong, W., Shen, K., and Feng, W. (2024). The structure and assembly of the hetero-octameric BLOC-one-related complex. Structure S0969– 2126(24)00536.

40. Li, W., Zhang, Q., Oiso, N., Novak, E.K., Gautam, R., O’Brien, E.P., Tinsley, C.L., Blake, D.J., Spritz, R.A., Copeland, N.G., et al. (2003). Hermansky-Pudlak syndrome type 7 (HPS-7) results from mutant dysbindin, a member of the biogenesis of lysosome-related organelles complex 1 (BLOC-1). Nat Genet 35, 84–89.

41. Morgan, N.V., Pasha, S., Johnson, C.A., Ainsworth, J.R., Eady, R.A., Dawood, B., McKeown, C., Trembath, R.C., Wilde, J., Watson, S.P., et al. (2006). A germline mutation in BLOC1S3/reduced pigmentation causes a novel variant of Hermansky-Pudlak syndrome (HPS8). Am J Hum Genet 78, 160–166.

42. Badolato, R., Prandini, A., Caracciolo, S., Colombo, F., Tabellini, G., Giacomelli, M., Cantarini, M.E., Pession, A., Bell, C.J., Dinwiddie, D.L., et al. (2012). Exome sequencing reveals a pallidin mutation in a Hermansky-Pudlak-like primary immunodeficiency syndrome. Blood 119, 3185–3187.

43. Cullinane, A.R., Curry, J.A., Golas, G., Pan, J., Carmona-Rivera, C., Hess, R.A., White, J.G., Huizing, M., and Gahl, W.A. (2012). A BLOC-1 mutation screen reveals a novel BLOC1S3 mutation in Hermansky-Pudlak Syndrome type 8. Pigment Cell Melanoma Res 25, 584–591.

44. Lowe, G.C., Sánchez Guiu, I., Chapman, O., Rivera, J., Lordkipanidzé, M., Dovlatova, N., Wilde, J., Watson, S.P., Morgan, N.V., and UK, G.A.P.P.C. (2013). Microsatellite markers as a rapid approach for autozygosity mapping in Hermansky-Pudlak syndrome: identification of the second HPS7 mutation in a patient presenting late in life. Thromb Haemost 109, 766–768.

45. Yousaf, S., Shahzad, M., Kausar, T., Sheikh, S.A., Tariq, N., Shabbir, A.S., University, O.W.C.F.M.G., Ali, M., Waryah, A.M., Shaikh, R.S., et al. (2016). Identification and clinical characterization of Hermansky-Pudlak syndrome alleles in the Pakistani population. Pigment Cell Melanoma Res 29, 231–235.

46. Bryan, M.M., Tolman, N.J., Simon, K.L., Huizing, M., Hufnagel, R.B., Brooks, B.P., Speransky, V., Mullikin, J.C., Gahl, W.A., Malicdan, M.C.V., et al. (2017). Clinical and molecular phenotyping of a child with Hermansky-Pudlak syndrome-7, an uncommon genetic type of HPS. Mol Genet Metab 120, 378–383.

47. Lasseaux, E., Plaisant, C., Michaud, V., Pennamen, P., Trimouille, A., Gaston, L., Monfermé, S., Lacombe, D., Rooryck, C., Morice-Picard, F., et al. (2018). Molecular characterization of a series of 990 index patients with albinism. Pigment Cell Melanoma Res 31, 466–474.

48. Bastida, J.M., Morais, S., Palma-Barqueros, V., Benito, R., Bermejo, N., Karkucak, M., Trapero-Marugan, M., Bohdan, N., Pereira, M., Marin-Quilez, A., et al. (2019). Identification of novel variants in ten patients with Hermansky-Pudlak syndrome by high-throughput sequencing. Ann Med 51, 141–148.

49. Okamura, K., Abe, Y., Araki, Y., Wakamatsu, K., Seishima, M., Umetsu, T., Kato, A., Kawaguchi, M., Hayashi, M., Hozumi, Y., et al. (2018). Characterization of melanosomes and melanin in Japanese patients with Hermansky-Pudlak syndrome types 1, 4, 6, and 9. Pigment Cell Melanoma Res 31, 267–276.

50. Pennamen, P., Le, L., Tingaud-Sequeira, A., Fiore, M., Bauters, A., Van Duong Béatrice, N., Coste, V., Bordet, J.C., Plaisant, C., Diallo, M., et al. (2020). BLOC1S5 pathogenic variants cause a new type of Hermansky-Pudlak syndrome. Genet Med 22, 1613–1622.

51. Boeckelmann, D., Wolter, M., Käsmann-Kellner, B., Koehler, U., Schieber-Nakamura, L., and Zieger, B. (2021). A Novel Likely Pathogenic Variant in the BLOC1S5 Gene Associated with Hermansky-Pudlak Syndrome Type 11 and an Overview of Human BLOC-1 Deficiencies. Cells 10, 2630.

52. Michaud, V., Fiore, M., Coste, V., Huguenin, Y., Bordet, J.C., Plaisant, C., Lasseaux, E., Morice-Picard, F., and Arveiler, B. (2021). A new case with Hermansky-Pudlak syndrome type 9, a rare cause of syndromic albinism with severe defect of platelets dense bodies. Platelets 32, 420–423.

53. Liu, T., Yuan, Y., Bai, D., Yao, X., Zhang, T., Huang, Q., Qi, Z., Yang, L., Yang, X., Li, W., et al. (2021). The first Hermansky-Pudlak syndrome type 9 patient with two novel variants in Chinese population. J Dermatol 48, 676–680.

54. Pennamen, P., Tingaud-Sequeira, A., Michaud, V., Morice-Picard, F., Plaisant, C., Vincent-Delorme, C., Giuliano, F., Azarnoush, S., Capri, Y., Marçon, C., et al. (2021). Novel variants in the BLOC1S3 gene in patients presenting a mild form of Hermansky-Pudlak syndrome. Pigment Cell Melanoma Res 34, 132–135.

55. Zhong, Z., Wu, Z., Zhang, J., and Chen, J. (2021). A novel BLOC1S5-related HPS-11 patient and zebrafish with bloc1s5 disruption. Pigment Cell Melanoma Res 34, 1112– 1119.

56. Zhang, A., He, X., Zhang, L., Yang, L., Woodman, P., and Li, W. (2014). Biogenesis of lysosome-related organelles complex-1 subunit 1 (BLOS1) interacts with sorting nexin 2 and the endosomal sorting complex required for transport-I (ESCRT-I) component TSG101 to mediate the sorting of epidermal growth factor receptor into endosomal compartments. J Biol Chem 289, 29180–29194.

57. Cheli, V.T., Daniels, R.W., Godoy, R., Hoyle, D.J., Kandachar, V., Starcevic, M., Martinez-Agosto, J.A., Poole, S., DiAntonio, A., Lloyd, V.K., et al. (2010). Genetic modifiers of abnormal organelle biogenesis in a Drosophila model of BLOC-1 deficiency. Hum Mol Genet 19, 861–878.

58. Chen, T., Song, G., Yang, H., Mao, L., Cui, Z., and Huang, K. (2018). Development of the Swimbladder Surfactant System and Biogenesis of Lysosome-Related Organelles Is Regulated by BLOS1 in Zebrafish. Genetics 208, 1131–1146.

59. Bultema, J.J., Ambrosio, A.L., Burek, C.L., and Di Pietro, S.M. (2012). BLOC-2, AP-3, and AP-1 proteins function in concert with Rab38 and Rab32 proteins to mediate protein trafficking to lysosome-related organelles. J Biol Chem 287, 19550–19563.

60. Niwa, S., Tao, L., Lu, S.Y., Liew, G.M., Feng, W., Nachury, M.V., and Shen, K. (2017). BORC regulates the axonal transport of synaptic vesicle precursors by activating ARL-8. Curr Biol 27, 2569–2578.e4.

61. Farías, G.G., Guardia, C.M., De Pace, R., Britt, D.J., and Bonifacino, J.S. (2017). BORC/kinesin-1 ensemble drives polarized transport of lysosomes into the axon. Proc Natl Acad Sci U S A 114, E2955–E2964.

62. Ferreirinha, F., Quattrini, A., Pirozzi, M., Valsecchi, V., Dina, G., Broccoli, V., Auricchio, A., Piemonte, F., Tozzi, G., Gaeta, L., et al. (2004). Axonal degeneration in paraplegin-deficient mice is associated with abnormal mitochondria and impairment of axonal transport. J Clin Invest 113, 231–242.

63. Denton, K.R., Lei, L., Grenier, J., Rodionov, V., Blackstone, C., and Li, X.J. (2014). Loss of spastin function results in disease-specific axonal defects in human pluripotent stem cell-based models of hereditary spastic paraplegia. Stem Cells 32, 414–423.

64. Thapa, D., Wu, K., Stoner, M.W., Xie, B., Zhang, M., Manning, J.R., Lu, Z., Li, J.H., Chen, Y., Gucek, M., et al. (2018). The protein acetylase GCN5L1 modulates hepatic fatty acid oxidation activity via acetylation of the mitochondrial β-oxidation enzyme HADHA. J Biol Chem 293, 17676–17684.

65. Thapa, D., Bugga, P., Mushala, B.A.S., Manning, J.R., Stoner, M.W., McMahon, B., Zeng, X., Cantrell, P.S., Yates, N., Xie, B., et al. (2022). GCN5L1 impairs diastolic function in mice exposed to a high fat diet by restricting cardiac pyruvate oxidation. Physiol Rep 10, e15415.

66. Zhang, M., Feng, N., Peng, Z., Thapa, D., Stoner, M.W., Manning, J.R., McTiernan, C.F., Yang, X., Jurczak, M.J., Guimaraes, D., et al. (2023). Reduced acetylation of TFAM promotes bioenergetic dysfunction in the failing heart. iScience 26, 106942.

67. Stewart, J.E., Crawford, J.M., Mullen, W.E., Jacques, A., Stoner, M.W., Scott, I., and Thapa, D. (2024). Cardiomyocyte-specific deletion of GCN5L1 reduces lysine acetylation and attenuates diastolic dysfunction in aged mice by improving cardiac fatty acid oxidation. Biochem J 481, 423–436.

68. Yang, C., Yang, Y., Hu, X., Tang, Q., Zhang, J., Zhang, P., Lu, X., Xu, J., Li, S., Dong, Z., et al. (2024). Loss of GCN5L1 exacerbates damage in alcoholic liver disease through ferroptosis activation. Liver Int 44, 1924–1936.

69. Williamson, C.D., Guardia, C.M., De Pace, R., Bonifacino, J.S., and Saric, A. (2022). Measurement of lysosome positioning by shell analysis and line scan. Methods Mol Biol 2473, 285–306.

70. Lord, S.J., Velle, K.B., Mullins, R.D., and Fritz-Laylin, L.K. (2020). SuperPlots: Communicating reproducibility and variability in cell biology. J Cell Biol 219, e202001064.

